# Single-fiber morphometry and spatial transcriptomics reveal selective oxidative muscle fiber atrophy in non-metastatic breast cancer

**DOI:** 10.64898/2026.05.11.26351978

**Authors:** Alan D. Mizener, Stuart A. Clayton, Alexa L. Bostic, Isabelle A. Oberhauser, Hannah E. Wilson, Marcella A. Whetsell, Hannah Hazard-Jenkins, Jessica F. Partin, Emidio E. Pistilli

## Abstract

Cancer-related fatigue is the most common and persistent symptom in breast cancer, with fatigue reported up to 10 years post-diagnosis. Unlike many cancers, fatigue in breast cancer often arises during early-stage disease in the absence of cachexia. While many factors contribute to fatigue, the direct contribution of cancer-associated skeletal muscle pathology remains poorly understood. Here we analyzed pectoralis major muscle biopsies from individuals with non-metastatic breast cancer and non-cancer controls using single-fiber morphometry and spatial transcriptomics. We identified fiber-type-specific structural alterations and spatially localized transcriptional reprogramming within the muscle microenvironment. Single-fiber morphometry revealed selective atrophy of oxidative type I and type IIa muscle fibers, while glycolytic type IIx fibers were relatively preserved. Concordant spatial transcriptomic profiling revealed suppression of oxidative metabolic programs, evidence of mitochondrial dysfunction, and spatially localized catabolic signaling originating from intramuscular adipocytes. This study introduces an integrated framework for profiling skeletal muscle architecture and spatially localized gene expression in surgically obtained muscle biopsies and represents the first application of spatial transcriptomics to human skeletal muscle from individuals with cancer. These findings demonstrate structural and metabolic remodeling of skeletal muscle in non-metastatic breast cancer and suggest targeting muscle metabolism represents a promising therapeutic strategy for cancer-related fatigue.

## Introduction

As breast cancer survival improves, long-term symptom burden has emerged as a defining survivorship challenge, with fatigue being among the most prevalent and persistent sequelae. A meta-analysis of 27 studies involving more than 12,000 participants estimated breast cancer severe fatigue prevalence at 26.9%.^1^ Reports of fatigue prevalence during active treatment are as high as 84%,^2^ while long-term studies estimate prevalence between 38-41% at 2-5 years^3,4^ and 27%^5^ at 10 years post-diagnosis. Improvements in survival have transformed breast cancer into a chronic condition for many individuals, amplifying the adverse long-term impact of persistent fatigue on quality of life and overall health.^6,7^ However, despite its prevalence and impact, there are no approved therapies for cancer-related fatigue. The persistent and multifactorial nature of breast cancer-related fatigue highlights a critical need to define its underlying pathophysiology to inform the development of effective, targeted interventions.

Current screening paradigms for muscle dysfunction in cancer rely heavily on measures of body weight loss and clinical classifications for cachexia.^8^ Yet, despite the relatively high prevalence of fatigue, breast cancer has one of the lowest rates of diagnosed cachexia at 0.9%.^9^ While cachexia definitions are designed to capture overt skeletal muscle wasting, clinically occult muscle pathology may substantially contribute to the development of fatigue in breast cancer. The etiology of cancer-related fatigue is multifactorial, including biological, medical, psychosocial, and demographic factors.^10–12^ However, previous work on breast cancer-induced skeletal muscle fatigue has demonstrated that breast cancer xenografts alone are sufficient to drive measurable contractile deficits and fatigue-associated transcriptional changes in the skeletal muscle of implanted mice.^13^ The same transcriptional changes are also reliably observed in individuals with breast cancer, regardless of treatment history or tumor molecular subtype.^14^ These pathologic alterations have previously been linked to the downregulation of the transcription factor peroxisome proliferator-activated receptor gamma (PPARG),^13^ a central regulator of lipolysis and mitochondrial biogenesis.^15^ Subsequent studies have shown that the use of PPARG agonists can partially rescue many fatigue-associated transcriptional changes,^16–18^ but fail to restore observed contractile deficits.^16,18^ One possible explanation for this apparent inconsistency between transcriptional and contractile function is that previous work used bulk sequencing approaches, which cannot resolve fiber-type-specific effects and non-myofiber contributions to fatigue, underscoring the importance of resolving fatigue-associated transcriptional changes by muscle cell type.

We hypothesized that subclinical skeletal muscle atrophy occurs even in early-stage breast cancer, and that transcriptional and metabolic remodeling of various cellular populations within the muscle microenvironment contribute to this occult atrophy and the development of fatigue. To test this hypothesis, we analyzed pectoralis major muscle biopsies from individuals undergoing breast surgery for non-metastatic breast cancer and non-malignant indications. Integrated single-fiber morphometry and spatial transcriptomic (ST) profiling revealed selective atrophy of oxidative type I and type IIa muscle fibers in the breast cancer cohort, accompanied by transcriptional suppression of oxidative metabolism and activation of catabolic signaling programs within these same muscle fiber populations. Interestingly, intramuscular adipocytes were the source of previously reported PPARG gene network suppression^13^ and also exhibited elevated expression of catabolic adipokine transcripts. Enrichment of corresponding receptor transcripts in adjacent muscle supports the possibility of catabolic paracrine signaling interactions originating from dysregulated intramuscular adipocytes within the muscle microenvironment. Collectively, these findings demonstrate that clinically occult skeletal muscle pathology is present in non-metastatic breast cancer prior to the development of overt cancer cachexia and suggest coordinated metabolic and catabolic reprogramming as an early feature of the disease. These alterations may contribute to the high prevalence of fatigue in the absence of body weight loss observed in individuals with breast cancer and highlight potential avenues for muscle-targeted therapeutic intervention.

## Results

Pectoralis major muscle biopsies were obtained from individuals with breast cancer (n = 16) and individuals undergoing breast surgery for non-malignant indications (n = 4). The clinical characteristics of the study participants are summarized in Table 1. The two cohorts were comparable with respect to age, body mass index (BMI), and body fat percentage. The breast cancer cohort consisted of individuals with non-metastatic disease (American Joint Committee on Cancer [AJCC] clinical anatomic stage I-III) and included an approximately even distribution across molecular subtypes, including luminal, HER2-positive, triple-positive, and triple-negative disease. Neoadjuvant chemotherapy was administered to 10 of the 16 participants in the breast cancer cohort prior to muscle biopsy collection, with most HER2-positive and triple-positive individuals receiving a HER2-targeted TCHP (docetaxel, carboplatin, trastuzumab, and pertuzumab) regimen and most triple-negative individuals receiving a KEYNOTE-522-based regimen,^19^ consistent with current standards of care. None of the luminal participants received neoadjuvant chemotherapy. Notably, there was no clinical evidence of systemic wasting in the breast cancer cohort, suggesting that any observed muscle alterations occur in the absence of overt cachexia. Spatial transcriptomics (ST) analyses were performed on muscle biopsies from two early-stage hormone receptor-positive breast cancer participants without prior chemotherapy exposure, along with two non-cancer controls, while other assays were conducted across the full cohort.

**Table 1.** – Clinical characteristics for the non-cancer and breast cancer cohorts.

|  | Non-cancer<br>(n = 4) | Breast cancer (n = 16) | P<br>value |
| --- | --- | --- | --- |
| Age | 48 (40–65) | 53 (25–70) | 0.47 |
| BMI (kg/m<sup>2</sup>) | 28.8 (20–35) | 31.1 (20–60) | 0.58 |
| Body fat (%) | 39.5 (30–45) | 35.9 (25–55) | 0.37 |
| FACIT-F fatigue subscale score | 32 (15–50) | 31 (5–50) | 0.90 |
| AJCC clinical anatomic stage | — | I: 4 (25%); II: 7 (44%); III: 5 (31%) | — |
| Molecular subtype | — | Luminal: 4 (25%); HER2: 5 (31%); TP: 4 (25%); TNBC: 3 (19%) | — |
| NACT | — | 10 (63%) | — |
| NACT regimen | — | None: 6 (38%); TCHP: 5 (31%); THP: 2 (13%); KN-522: 3 (19%) | — |

BMI, body mass index; FACIT-F, Functional Assessment of Chronic Illness Therapy–Fatigue;
AJCC, American Joint Committee on Cancer; NACT, neoadjuvant chemotherapy; Luminal
(hormone receptor-positive [HR+]/human epidermal growth factor receptor 2-negative [HER2–]);
HER2 (HR–/HER2+); TP (HR+/HER2+); TNBC (HR–/HER2–); TCHP,
docetaxel/carboplatin/trastuzumab/pertuzumab; THP, docetaxel/trastuzumab/pertuzumab; KN-522, pembrolizumab/paclitaxel/carboplatin followed by
pembrolizumab/doxorubicin/cyclophosphamide. Continuous variables are presented as mean
(range). Ranges for continuous clinical variables were rounded to the nearest multiple of five to
reduce the risk of participant re-identification. Categorical variables are presented as count (%).
P values were calculated using two-sided Welch's t-tests. Percentages may not sum to 100%
due to rounding.

Participant-reported fatigue, assessed using the Functional Assessment of Chronic Illness Therapy–Fatigue (FACIT-F) questionnaire,^20^ did not differ significantly between the non-cancer and breast cancer cohorts (Table 1). Because FACIT-F measures subjective perceptions of fatigue rather than objective skeletal muscle function, comparisons between the breast cancer and non-cancer populations may be influenced by differences in baseline expectations of physical function.^21^ Clinical characteristics and assay utilization for each biopsy are provided in Supplementary Table 1 and Supplementary Table 2, respectively.

### Oxidative muscle fibers exhibit selective atrophy in non-metastatic breast cancer

To determine if subclinical skeletal muscle atrophy occurs with breast cancer, we performed immunofluorescent muscle fiber-typing on pectoralis major biopsies obtained from non-metastatic breast cancer and non-cancer surgical cohorts (Fig. 1A). Muscle fibers were segmented from the laminin channel and classified using a supervised machine-learning pipeline (Fig. 1B). Type I, type IIa, and type IIx fiber types were then assigned to high-quality cross-sectional fibers based on MYH7, MYH2, and MYH1 channel features, respectively (Fig. 1C). Model validation demonstrated high discriminative performance across all segmentation classes and muscle fiber types (Supplementary Fig. 1).

**Figure 1.**
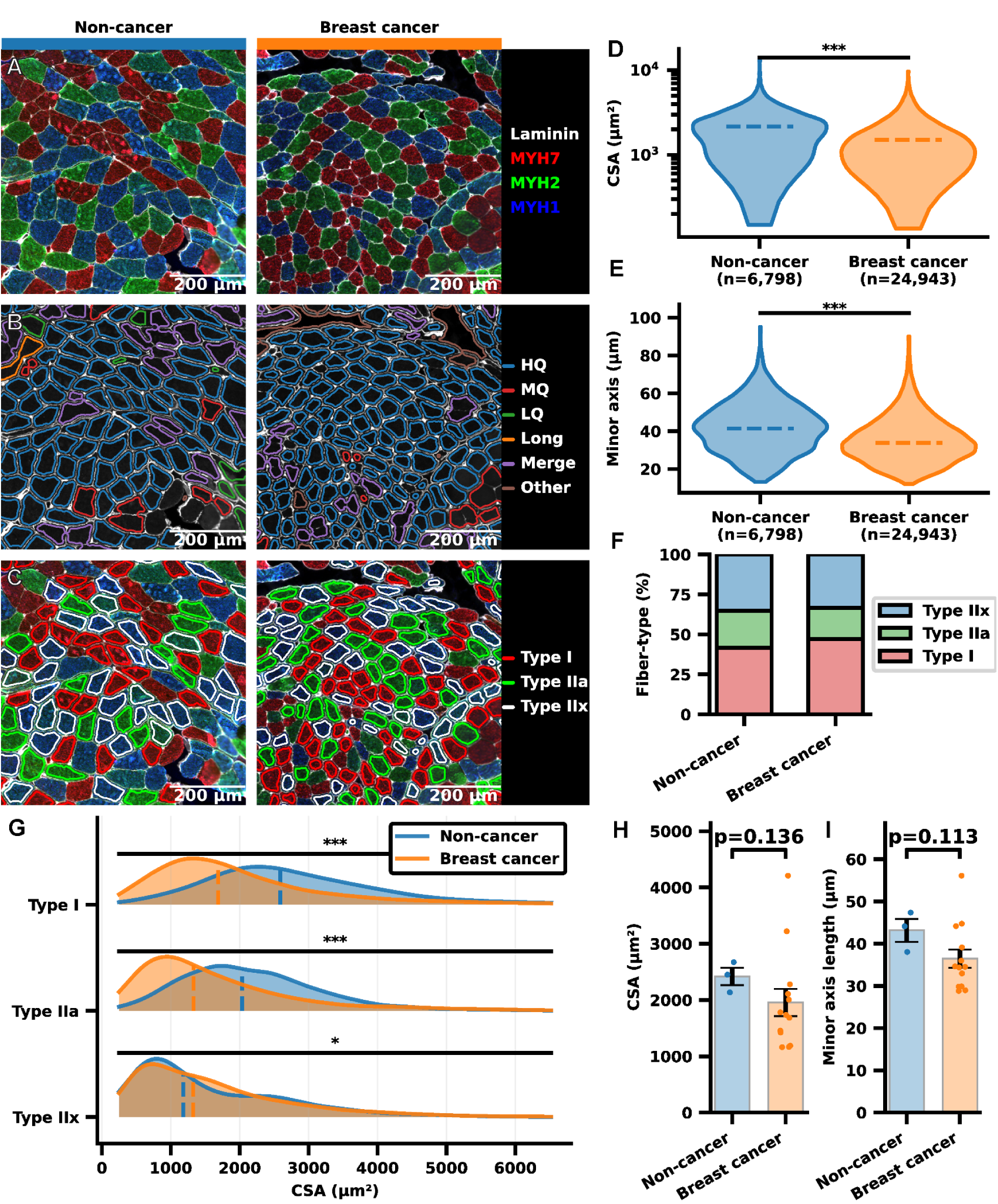
– Single fiber morphometry reveals selective atrophy of oxidative muscle fibers in non-metastatic breast cancer. **A** Representative cross-sectional images of pectoralis major biopsies with immunofluorescent (IF) fiber-type staining. **B** Representative segmentation classifications from the machine-learning (ML) classifier. HQ=high-quality cross-sectional muscle fiber; MQ=medium-quality cross-sectional muscle fiber; LQ=low-quality muscle fiber; Long=longitudinal muscle fiber; Merge=merged muscle fiber; Other=other structure. **C** Representative fiber-typing classifications from the ML classifier. **D-E** Comparison of muscle fiber cross-sectional area (CSA; **D**) and minor axis length (**E**). n = single muscle fibers. Dashed lines represent medians. **F** Proportion of muscle fiber types split by cohort. **G** Stratification of muscle fiber CSA by fiber type. Dashed lines represent medians. **H-I** Individual-level comparisons of CSA (**H**) and minor axis length (**I**). Sections with >1000 high-quality fibers were used to compute per-individual means. n = individual study participants. Dots represent individual participant means. Error bars represent means ± standard error of the mean. **D-I** Comparisons between cohorts were performed using a two-tailed Welch’s t-test. *P < 0.05; **P < 0.01; ***P < 0.001.

In aggregate, muscle fibers from the breast cancer cohort exhibited significantly smaller cross-sectional areas (Fig. 1D) and shorter minor axis lengths (Fig. 1E). Stratification by AJCC clinical anatomic stage revealed that this reduction was consistent across stages (Supplementary Fig. 2D), indicating that muscle fiber atrophy occurs early in disease progression. The presence of muscle fiber atrophy in individuals with early-stage breast cancer suggests that pathologic skeletal muscle remodeling is not restricted to locally advanced disease and may contribute to the high prevalence of fatigue reported in breast cancer. Fiber-type composition was preserved between cohorts (Fig. 1F), suggesting that the observed differences in muscle fiber size were not attributable to shifts in the fiber-type distribution.

Stratification by muscle fiber-type revealed selective atrophy of oxidative type I (−35%) and type IIa (−35%) fibers by median cross-sectional area, whereas glycolytic type IIx fibers were relatively preserved (Fig. 1G). Although type IIx fibers exhibited a small but statistically significant increase in median cross-sectional area (+12%), this effect was not accompanied by changes in minor axis length (Supplementary Fig. 2A), suggesting limited structural remodeling of type IIx fibers. These findings indicate a preferential vulnerability of oxidative muscle fibers in the setting of non-metastatic breast cancer.

To determine if muscle fiber atrophy was accompanied by alterations in intramuscular glycogen, we performed periodic acid-Schiff (PAS) staining of adjacent muscle sections. PAS reactivity did not differ qualitatively between cohorts (Supplementary Fig. 3), indicating that non-metastatic breast cancer is not associated with significant changes in intramuscular glycogen content.

To account for potential individual-level variability, we repeated these analyses on a per-participant basis after excluding sections with fewer than 1,000 high-quality cross-sectional muscle fibers. Directionally consistent reductions in cross-sectional area (Fig. 1H) and minor axis length (Fig. 1I) were observed in the breast cancer cohort, although these reductions were not statistically significant. Examination of clinical covariates identified one sample with an associated BMI that exceeded the upper Tukey outlier threshold (Q3+1.5xIQR=48.15 kg/m²). In a follow-up individual-level analysis excluding this sample, the reduction in aggregate muscle fiber cross-sectional area reached statistical significance (p=0.023; Supplementary Fig. 2B), and a similar trend was observed for minor axis length (p=0.066; Supplementary Fig. 2C).

Aggregate muscle fiber cross-sectional area was also positively correlated with FACIT-F fatigue subscale score in the breast cancer cohort (r=0.61, p=0.04; Supplementary Fig. 2G), suggesting that the FACIT-F may reflect underlying changes in muscle fiber morphology and could be developed as a non-invasive screening modality for the detection of early muscle fiber atrophy, warranting further investigation. Additional analyses examining the influence of AJCC clinical anatomic stage, breast cancer molecular subtype, neoadjuvant chemotherapy exposure, age, and BMI on aggregate muscle fiber cross-sectional area are shown in Supplementary Fig. 2.

Collectively, these data demonstrate that selective atrophy of oxidative type I and type IIa muscle fibers occurs in non-metastatic breast cancer, even in the absence of overt cachexia, while glycolytic type IIx fibers are comparatively preserved.

### Spatial transcriptomics resolves type I and type IIa transcriptional profiles

Given the selective reduction in oxidative muscle fiber size observed in breast cancer, we next sought to characterize tumor-associated transcriptional programs within the skeletal muscle in a minimally confounded clinical context. ST profiling was performed on adjacent pectoralis major biopsy sections from a subset of chemotherapy-naive, early-stage hormone receptor-positive cases (N=2 breast cancer; N=2 non-cancer). Following standard quality-control filtering and normalization (Supplementary Figs. 4, 5, and 6), unsupervised clustering resolved transcriptionally distinct muscle fiber and non-myofiber spot populations (Fig. 2A). Spatial projection of the annotated clusters revealed anatomically coherent localization within each tissue, with representative non-cancer and breast cancer sections shown (Fig. 2B). Cluster proportions were comparable between cohorts (Fig. 2C), indicating preservation of overall cellular composition in early-stage breast cancer.

**Figure 2.**
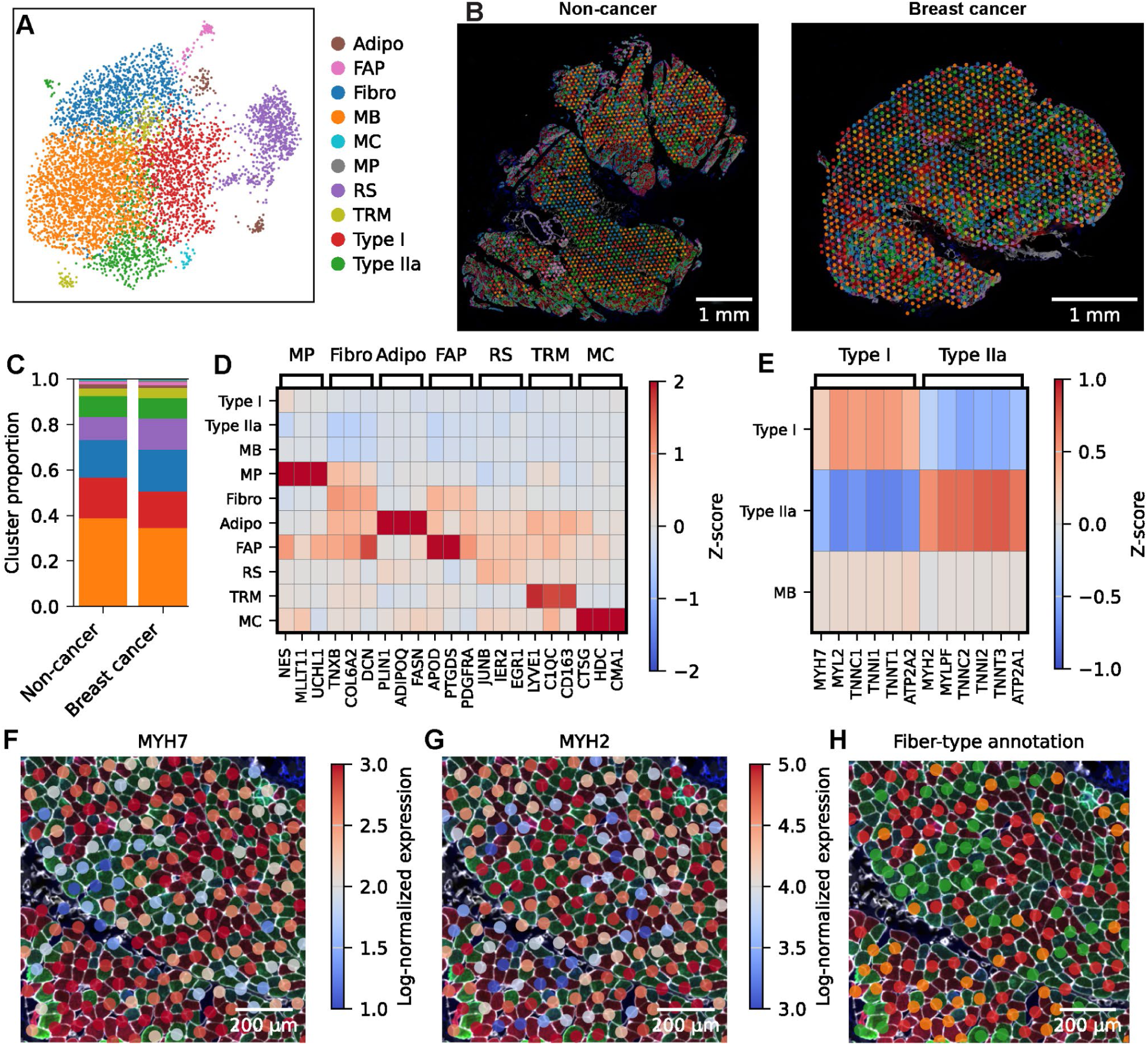
– Spatial transcriptomics resolves type I and type IIa muscle fiber transcriptional profiles. **A** Uniform manifold approximation and projection (UMAP) of spot-types identified by spatial transcriptomics from pectoralis major muscle biopsies. n=single ST spots. Adipo=adipocytes; FAP=fibro-adipogenic progenitors; Fibro=fibroblasts; MB=muscle background; MC=mast cells; MP=myogenic progenitors; RS=reactive stroma; TRM=tissue-resident macrophages; Type I=type I muscle fibers; Type IIa=type IIa muscle fibers. **B** Representative spatial overlays of spot annotations. **C** Spot-type annotation proportions for the non-cancer and breast cancer cohorts. **D** Marker gene matrix for muscle stromal cell marker genes. **E** Marker gene matrix for muscle fiber-type marker genes. **F** Representative spatial overlay of MYH7 transcript expression on IF fiber-typed muscle. **G** Representative spatial overlay of MYH2 transcript expression on IF fiber-typed muscle. **H** Representative spatial overlay of spot-type annotations on IF fiber-typed muscle. Non-myofiber annotations not shown.

Canonical and differentially expressed marker genes demonstrated strong specificity for annotated non-myofiber populations, with minimal intervening expression across other clusters (Fig. 2D). In contrast, myofiber marker genes were broadly expressed across all spots, resulting in a unimodal marker expression distribution. Fiber-type-specific populations were therefore defined by relative enrichment of type I or type IIa transcriptional signatures rather than by binary marker restriction, alongside a “muscle background” (MB) population exhibiting mixed expression of both markers without a clear fiber-type predominance (Fig. 2E).

Spatial mapping of MYH7 and MYH2 expression onto aligned immunofluorescent fiber-typed sections demonstrated precise concordance with type I (Fig. 2F) and type IIa fibers (Fig. 2G), respectively, with myofiber spot annotations exhibiting a similar, robust pattern of co-localization (Fig. 2H). Consistent with the spatial resolution of the 10x Visium v2 assay, mixed-marker “muscle background” spots preferentially localized to muscle fiber-type boundaries (Fig. 2H). This population was included in aggregated muscle analyses but analyzed separately when assessing fiber-type specificity.

Collectively, these analyses establish ST as a robust platform for resolving fiber-type-specific transcriptional states and associated cellular populations within surgically obtained skeletal muscle biopsies.

### Transcriptional suppression of mitochondrial oxidative metabolism in type I and type IIa fibers

To assess fiber-type-specific transcriptional alterations in early-stage breast cancer, we compared type I, type IIa, and muscle background populations between cohorts using differential gene expression analysis. Gene set enrichment analysis (GSEA) revealed oxidative phosphorylation as the top downregulated pathway in breast cancer across all myofiber populations (Fig. 3A). The magnitude of pathway suppression was greatest in type I fibers, followed by muscle background and type IIa populations, consistent with their relative reliance on oxidative metabolism.^22^ Notably, transcriptional suppression of oxidative phosphorylation is consistent with our prior bulk-RNA-sequencing analysis of pectoralis major muscle biopsies from a larger breast cancer cohort (N=10 non-cancer, N=33 breast cancer), which similarly identified mitochondrial oxidative metabolism as a prominent downregulated transcriptional program.^14^

**Figure 3.**
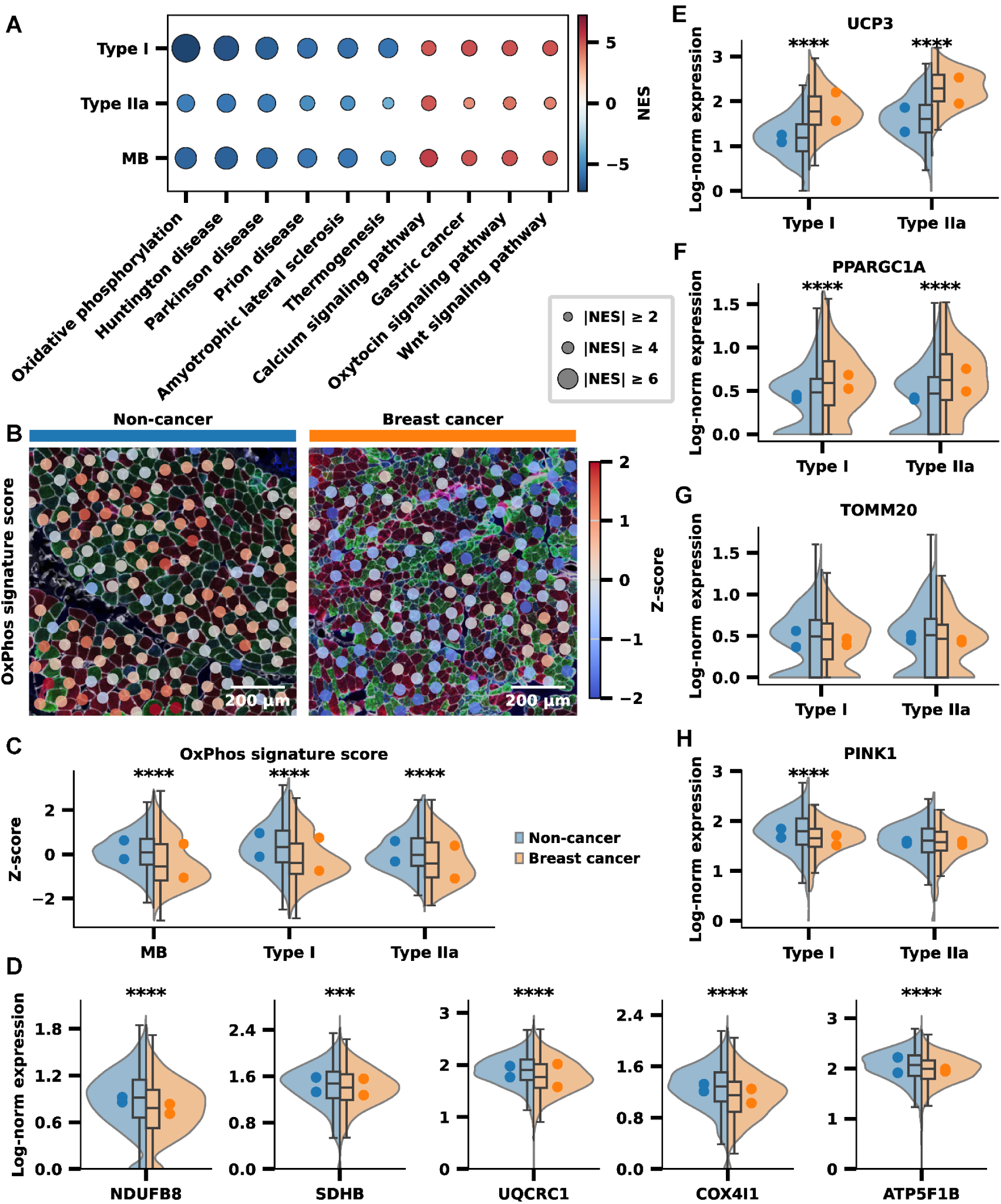
– Differential expression analysis reveals transcriptional suppression of oxidative metabolism in type I and type IIa muscle fibers from early-stage breast cancer. **A** Dot plot depicting the top 10 dysregulated KEGG 2021 Human gene sets. Genes were ranked based on signed t-statistics. Positive normalized enrichment scores (NES) represent enrichment in the breast cancer cohort, while negative NES represent enrichment in the non-cancer cohort. All depicted gene sets had an FDR-adjusted P value < 0.05. Type I=type I muscle fibers; Type IIa=type IIa muscle fibers; MB=muscle background; **B** Spatial overlay of normalized UCell KEGG 2021 Human Oxidative Phosphorylation signature scores. Non-myofiber annotations not shown. **C** Quantification of normalized UCell KEGG 2021 Human Oxidative Phosphorylation signature scores stratified by myofiber spot-type annotation. **D** Comparison of representative oxidative phosphorylation transcript expression in all aggregated myofiber spot types (type I, type IIa, MB), representing each of the electron transport chain complexes. Complex I=NDUFB8, Complex II=SDHB, Complex III=UQCRC1, Complex IV=COX4I1, Complex V=ATP5F1B. **E-H** Mitochondrial gene expression in type I and type IIa annotated spots for select dysregulated transcripts: UCP3 (**E**), PPARGC1A (**F**), TOMM20 (**G**), PINK1 (**H**). **C-H** Comparisons between cohorts were performed using a two-tailed Mann-Whitney U test with a Benjamini-Hochberg false discovery rate (FDR) correction. n = ST spots. Violin plots show spot-level distributions; boxplots indicate median and interquartile range (IQR), with whiskers extending to 1.5 x IQR; dots represent individual participant means. *P < 0.05; **P < 0.01; ***P < 0.001; ****P < 0.0001.

To further interrogate oxidative metabolic suppression in a spatial context, we computed per-spot oxidative phosphorylation signature scores using UCell^23,24^ and projected these scores onto tissue sections (Fig. 3B). Per-spot scoring recapitulated the prior GSEA findings, with all myofiber populations exhibiting significantly lower signature scores in the breast cancer cohort compared to non-cancer controls, with a coherent hierarchy of effect sizes (Fig. 3C). Consistent with established muscle fiber biology, type I fibers within each section exhibited greater oxidative phosphorylation signature scores compared to type IIa fibers, reflecting their comparatively higher oxidative capacity.^22^

To determine if oxidative metabolic suppression reflected coordinated downregulation across the electron transport chain (ETC), we examined representative transcripts from Complexes I-V in aggregated myofiber spots. Transcripts corresponding to each ETC complex were significantly downregulated in the breast cancer cohort compared to non-cancer controls (Fig. 3D), consistent with broad transcriptional suppression of mitochondrial respiratory machinery.

Among the most significantly upregulated transcripts in the breast cancer cohort was uncoupling protein 3 (UCP3), a transcript implicated in modulating mitochondrial coupling efficiency. UCP3 expression was significantly increased in both type I and type IIa myofiber populations (Fig. 3E). Induction of UCP3 in skeletal muscle has been associated with oxidative stress responses and shifts toward increased fatty acid utilization,^25^ suggesting altered skeletal muscle mitochondrial coupling and substrate handling in early-stage breast cancer.

To determine if mitochondrial regulatory programs governing biogenesis, content, and quality control were altered in early-stage breast cancer, we examined representative transcripts within each axis. Expression of peroxisome proliferator-activated receptor gamma coactivator 1-alpha (PPARGC1A), a master regulator of mitochondrial biogenesis,^26^ was significantly increased in both type I and type IIa myofiber populations in the breast cancer cohort (Fig. 3F), consistent with increased biogenic signaling. Despite increased PPARGC1A expression, transcripts associated with mitochondrial content, including translocase of outer mitochondrial membrane 20 (TOMM20),^27^ were not significantly altered in either fiber type (Fig. 3G). In contrast, PTEN-induced kinase 1 (PINK1), a regulator of mitochondrial quality control,^28^ was significantly downregulated in type I fibers but unchanged in type IIa fibers (Fig. 3H), suggesting disrupted mitochondrial damage surveillance, particularly in type I fibers. Collectively, these data demonstrate coordinated transcriptional remodeling of mitochondrial respiratory, regulatory, and quality-control programs in oxidative myofibers from individuals with early-stage breast cancer.

### Intramuscular adipocytes exhibit PPARG transcriptional network suppression and emerge as a potential paracrine source of catabolic adipokines

Given prior work implicating reduced peroxisome proliferator-activated receptor gamma (PPARG) signaling in fatigue-associated transcriptional remodeling of early-stage breast cancer skeletal muscle,^13,14,16–18^ we examined the spatial distribution of PPARG expression within the skeletal muscle microenvironment. Spatial projection of PPARG expression revealed sparse transcript detection, with a minority of spots containing detectable reads (Fig. 4A). PPARG transcripts were predominantly detected in adipocyte-annotated spots, whereas myofiber spots minimally contributed to overall tissue expression (Fig. 4B).

**Figure 4.**
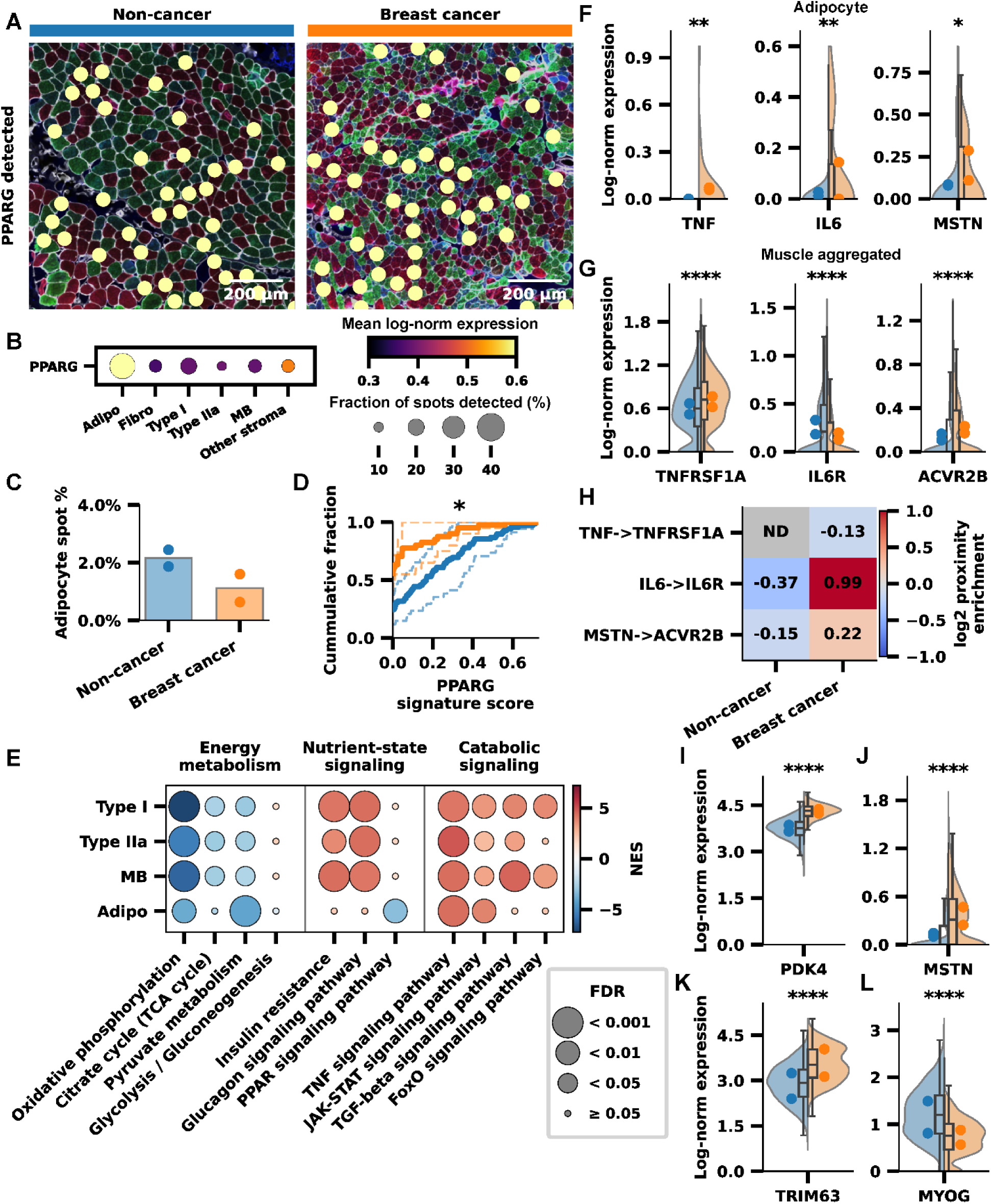
– Intramuscular adipocytes serve as a plausible source of catabolic signaling within the muscle microenvironment. **A** Representative spatial overlays identifying where PPARG transcripts were detected within skeletal muscle. **B** Dot plot depicting PPARG expression across spot-type annotations. Adipo=adipocytes; Fibro=fibroblasts; Type I=type I muscle fibers; Type IIa=type IIa muscle fibers; MB=muscle background; Other Stroma=all other annotations. **C** Bar plot depicting the percentage of tissue-covered spots annotated as adipocytes. Points represent individual section percentages. **D** Cumulative distribution function plot depicting the distribution of UCell PPARG signature scores in adipocyte-annotated spots. Solid lines represent cohort mean distributions. Dashed lines represent individual section distributions. Comparisons between cohorts were performed using a two-tailed Mann-Whitney U test with a Benjamini-Hochberg false discovery rate (FDR) correction. n = ST spots. *P < 0.05. **E** Dot plot depicting gene set enrichment results for curated KEGG 2021 Human gene sets grouped into biologic themes. Genes were ranked based on signed t-statistics. Positive normalized enrichment scores (NES) represent enrichment in the breast cancer cohort, while negative NES represent enrichment in the non-cancer cohort. Type I=type I muscle fibers; Type IIa=type IIa muscle fibers; MB=muscle background; Adipo=adipocytes. **F** Expression of literature-defined catabolic adipokines in adipocyte-annotated spots. **G** Expression of literature-defined receptors for catabolic adipokines in all aggregated myofiber spots (Type I, Type IIa, and MB). **H** Heatmap depicting log2-transformed proximity enrichment scores for literature-defined catabolic adipokine ligand-receptor pairs signaling from adipocyte-annotated spots to any myofiber-annotated spot (Type I, Type IIa, MB). Positive values indicate enriched spatial proximity. **I-L** Expression of select transcripts involved in substrate utilization and muscle fiber atrophy in all aggregated myofiber spots (Type I, Type IIa, MB). **F-G, I-L** Comparisons between cohorts were performed using a two-tailed Mann-Whitney U test with a Benjamini-Hochberg false discovery rate (FDR) correction. n = ST spots. Violin plots show spot-level distributions; boxplots indicate median and interquartile range (IQR), with whiskers extending to 1.5 x IQR; dots represent individual participant means. *P < 0.05; **P < 0.01; ***P < 0.001; ****P < 0.0001.

We next asked whether previously observed suppression of the PPARG transcriptional network could be explained by depletion of intramuscular adipocytes within the skeletal muscle microenvironment of early-stage breast cancer. Comparison of adipocyte-annotated spot proportions revealed a decrease in adipocyte abundance from 2.1% in non-cancer to 1.1% in the breast cancer cohort, representing a 48% relative reduction in adipocyte abundance (Fig. 4C), suggesting that depletion of intramuscular adipocytes may contribute to reduced PPARG transcriptional network expression. To determine if intramuscular adipocytes also exhibit cell-intrinsic reductions in expression of the PPARG transcriptional network, we computed single-spot PPARG signature scores using a published list of validated PPARG target genes.^13^ Cumulative distribution function (CDF) analysis revealed that a greater proportion of adipocyte-annotated spots in the breast cancer cohort exhibited a PPARG signature score of zero, and PPARG signature scores among spots with non-zero values were significantly lower compared to non-cancer controls (Fig. 4D), suggesting that cell-intrinsic downregulation of the PPARG transcriptional network by intramuscular adipocytes also contributes to reduced overall PPARG transcriptional activity.

To contextualize these adipocyte-localized transcriptional alterations within the broader skeletal muscle microenvironment, we next examined pathway-level transcriptional changes across myofiber and adipocyte populations. GSEA comparing breast cancer and non-cancer cohorts revealed coordinated alterations in pathways governing energy metabolism, nutrient-state signaling, and catabolic signaling across spatially resolved cell types within the muscle microenvironment (Fig. 4E). Consistent with earlier findings, myofiber populations exhibited broad suppression of mitochondrial metabolism, with oxidative phosphorylation, tricarboxylic acid (TCA) cycle, and pyruvate metabolism pathways significantly downregulated in the breast cancer cohort (Fig. 4E). In contrast, glycolytic pathways were largely unchanged across myofiber populations, consistent with the morphologic preservation of glycolytic type IIx fibers (Fig. 4E).

To determine if these shifts in energy metabolism were accompanied by changes in nutrient-state signaling, we next examined insulin resistance, glucagon signaling, and PPAR signaling pathways across the same spatially resolved spot populations. In the breast cancer cohort, myofiber populations exhibited significantly elevated insulin resistance and glucagon signaling, while suppression of PPAR signaling localized exclusively to intramuscular adipocytes (Fig. 4E), consistent with the adipocyte-restricted PPARG expression pattern observed in earlier analyses and suggesting broader metabolic dysregulation within the skeletal muscle microenvironment.

We also hypothesized that early activation of programs with established catabolic functions in skeletal muscle, including TNF, JAK-STAT (IL-6), TGF-beta (Myostatin), and FoxO (Atrogin-1), would accompany selective oxidative muscle fiber atrophy as described earlier. TNF, JAK-STAT, and TGF-beta signaling were significantly elevated across all myofiber populations in the breast cancer cohort, while FoxO signaling was significantly elevated primarily in the more oxidative type I-enriched and muscle background populations (Fig. 4E), consistent with activation of multiple catabolic transcriptional programs in oxidative muscle fibers that were previously identified as selectively atrophic. Notably, intramuscular adipocytes also exhibited elevated TNF and JAK-STAT signaling (Fig. 4E), suggesting that inflammatory signaling axes may also contribute to adipocyte transcriptional remodeling in early-stage breast cancer.

We next investigated whether intramuscular adipocytes could serve as a putative paracrine source of catabolic adipokines within the skeletal muscle microenvironment. To address this, we first examined the expression of established catabolic adipokines upstream of the transcriptional programs altered in myofibers. Tumor necrosis factor-alpha (TNF), interleukin-6 (IL-6), and myostatin (MSTN) were all significantly upregulated in intramuscular adipocytes from the breast cancer cohort (Fig. 4F).

We next assessed if each of the corresponding receptors were expressed by myofibers and whether their expression differed between cohorts. Tumor necrosis factor receptor superfamily member 1A (TNFRSF1A), the cognate receptor for TNF, and activin receptor type-2B (ACVR2B), the cognate receptor for MSTN, were both significantly upregulated in breast cancer myofiber populations (Fig. 4G). In contrast, interleukin-6 receptor (IL6R) was significantly downregulated in breast cancer (Fig. 4G).

To further evaluate the plausibility of adipocyte-to-myofiber paracrine signaling, we performed a ring-based spatial neighborhood proximity-enrichment analysis for each of the ligand-receptor pairs using adipocyte- and myofiber-annotated spots as the sending and receiving spot types, respectively. IL6-IL6R and MSTN-ACVR2B exhibited positive proximity enrichment (Fig. 4H), consistent with these axes representing plausible adipocyte-to-myofiber paracrine signaling mechanisms. In contrast, TNF-TNFRSF1A showed negative proximity enrichment (Fig. 4H), suggesting that TNF signaling within myofibers may be driven by systemic or inflammatory sources.

To highlight representative myofiber gene expression changes associated with the pathway alterations and microenvironment signaling axes described above, we next examined differentially expressed transcripts associated with metabolic remodeling and catabolic signaling. Pyruvate dehydrogenase kinase 4 (PDK4), a key regulator of mitochondrial substrate utilization that inhibits pyruvate entry into the TCA cycle,^29^ was significantly upregulated in the breast cancer cohort (Fig. 4I). Upregulation of PDK4 is consistent with the observed suppression of oxidative metabolism and suggests altered energy substrate utilization by myofibers in the setting of breast cancer.^30^ In addition to the adipocyte-derived MSTN signaling described above, myofibers also exhibited increased expression of MSTN in the setting of early-stage breast cancer (Fig. 4J), raising the possibility that both paracrine and autocrine signaling mechanisms contribute to TGF-beta catabolic signaling within myofibers.

We then examined key transcriptional markers associated with muscle atrophy and development. Tripartite motif containing 63 (TRIM63), an established “atrogene” and E3 ubiquitin ligase responsible for myofibrillar protein degradation,^31^ was significantly upregulated in breast cancer myofibers (Fig. 4K). In contrast, myogenin (MYOG), a key myogenic transcription factor that governs skeletal muscle differentiation and contributes to adult myofiber growth and muscle stem cell homeostasis,^32^ was significantly downregulated in breast cancer myofibers (Fig. 4L). Together, these transcriptional changes are consistent with the activation of established muscle atrophy programs and support the selective atrophy of oxidative muscle fibers observed by single-fiber morphometry in non-metastatic breast cancer.

## Discussion

In this study, we identified previously unrecognized oxidative muscle fiber atrophy in non-cachectic individuals with non-metastatic breast cancer. This structural phenotype is accompanied by transcriptional evidence of metabolic reprogramming, including suppression of oxidative phosphorylation and activation of catabolic signaling pathways, even in individuals with early-stage disease and no prior chemotherapy exposure. Spatial analyses further localized previously reported PPARG suppression^13^ to intramuscular adipocytes, implicating adipocyte to muscle fiber signaling as a potential mediator of the observed remodeling. These findings were enabled by the novel integration of spatial transcriptomics (ST) with single-fiber morphometry in muscle biopsies obtained during clinically indicated tumor resection procedures. Together, these results support a model in which multiple stressors converge to drive metabolic and structural remodeling of skeletal muscle in non-metastatic breast cancer, potentially contributing to the high prevalence and persistence of fatigue reported in this population^1,5^ (Fig. 5).

**Figure 5.**
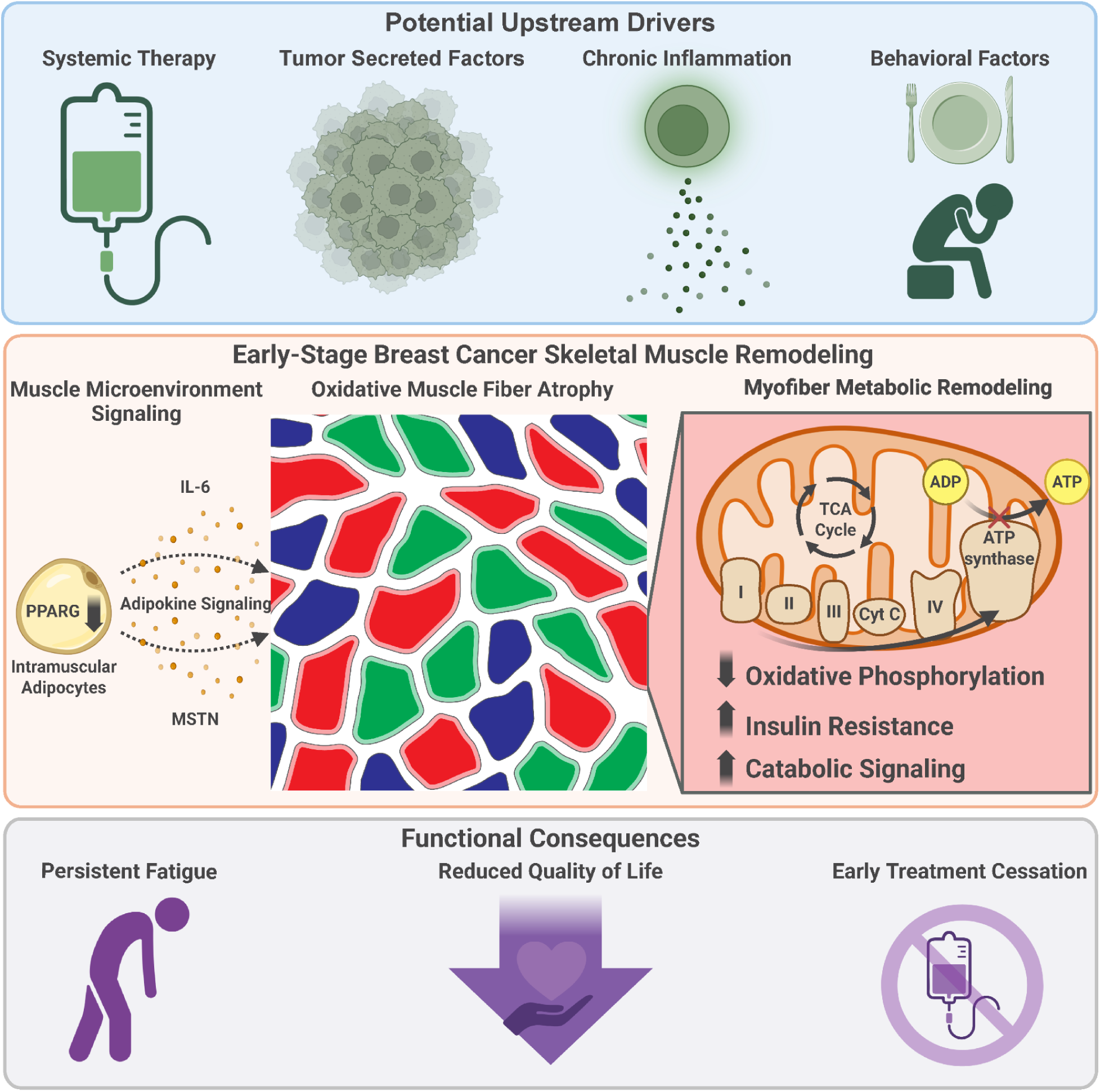
– Conceptual model of skeletal muscle remodeling in non-metastatic breast cancer.

Skeletal muscle dysfunction is increasingly recognized as a consequence of cancer, particularly in the context of advanced disease and cachexia.^33,34^ However, relatively little is known about skeletal muscle pathology in individuals with early-stage disease who do not exhibit overt cachexia. This knowledge gap has made the high prevalence and persistence of fatigue observed in cancers such as breast cancer^1,5^ difficult to reconcile with the absence of apparent wasting. In addition, most prior work has examined muscle at the tissue level,^14,35^ leaving fiber-type-specific structural and metabolic alterations, as well as the contributions of stromal cell populations such as intramuscular adipocytes, poorly characterized. The present study extends the literature by demonstrating that oxidative muscle fibers are preferentially susceptible to atrophy and metabolic reprogramming in non-metastatic breast cancer and by highlighting a potential role for intramuscular adipocytes as mediators of local catabolic signaling within skeletal muscle.

One of the most striking findings of the present study was the selective atrophy of oxidative (type I and type IIa) muscle fibers. This pattern contrasts with cancer cachexia, where glycolytic type II fibers, particularly type IIx, are generally reported to be most susceptible to atrophy.^36,37^ These findings suggest that skeletal muscle remodeling in non-metastatic breast cancer may arise through mechanisms distinct from those driving cancer cachexia. ST analyses in the present study, together with previously reported bulk RNA-sequencing,^14^ indicate that oxidative fiber atrophy occurs in conjunction with transcriptional suppression of oxidative phosphorylation. Given the heavy reliance of oxidative muscle fibers on mitochondrial respiration to sustain ATP production,^22^ suppression of oxidative pathways may disproportionately compromise their bioenergetic homeostasis. Additional transcriptional signals further suggest reduced mitochondrial coupling efficiency (UCP3 upregulation) and impaired mitochondrial quality control (PINK1 downregulation), consistent with broader mitochondrial dysfunction. Together, these findings suggest impaired mitochondrial metabolism may promote bioenergetic insufficiency within oxidative fibers, potentially contributing to their selective vulnerability in non-metastatic breast cancer. However, the mechanisms underlying this fiber-type specificity remain incompletely understood.

Another important advance of the present study is the spatial localization of PPARG suppression to intramuscular adipocytes and the clarification this provides regarding the role of PPARG signaling in breast cancer-associated skeletal muscle remodeling. We previously reported suppression of PPARG transcriptional networks in skeletal muscle from individuals with non-metastatic breast cancer using bulk RNA-sequencing approaches.^13^ However, the cellular origin of this signal could not be definitively resolved. Because PPARG, together with its coactivator PPARGC1A, is a key regulator of mitochondrial biogenesis and oxidative metabolism,^26^ we originally hypothesized that suppression of this pathway within muscle fibers might directly contribute to the oxidative metabolic defects observed in skeletal muscle from non-metastatic breast cancer.^14^ However, subsequent studies using pharmacologic PPARG agonists produced only partial improvements in muscle function.^16,18^ The ST analyses presented here clarify these findings by demonstrating that PPARG suppression is localized primarily to intramuscular adipocytes rather than myofibers. These adipocytes also emerge as a putative source of catabolic paracrine mediators within the muscle microenvironment, consistent with recent work suggesting non-muscle stromal cells may represent a dominant source of catabolic signaling in the skeletal muscle microenvironment during cancer cachexia.^38^ These results suggest that altered PPARG signaling in intramuscular adipocytes, rather than myofibers, may indirectly contribute to skeletal muscle remodeling in non-metastatic breast cancer through changes in the local metabolic and paracrine environment.

Although the present study identifies structural and metabolic remodeling of skeletal muscle as a feature of non-metastatic breast cancer, the upstream drivers of this phenotype remain incompletely understood. Multiple systemic stressors associated with cancer and its treatment may plausibly contribute to the skeletal muscle pathology observed here. These include effects of cytotoxic chemotherapy and endocrine therapy, both of which have been reported to adversely impact skeletal muscle.^39,40^ However, the biopsies profiled here using ST were obtained from individuals without prior systemic therapy exposure, suggesting that metabolic remodeling and atrophy can arise in the absence of treatment-derived stress. In such treatment-naive individuals, tumor-derived signals may play a more prominent role. Circulating cytokines, growth factors, and extracellular vesicles released by cancer cells are all capable of impacting skeletal muscle.^34,41,42^ Chronic inflammation from immune activation within the tumor microenvironment may also contribute to metabolic alterations and atrophy in skeletal muscle, particularly in the setting of cancer cachexia, where TNF-α is known to play a central role.^43^ Another plausible contributor is the “Warburg effect,” a phenomenon in which cancer cells preferentially rely on aerobic glycolysis to meet energy demands, resulting in excess consumption of available nutrients.^44^ Metabolic competition between the tumor and peripheral tissues could therefore be a plausible contributor to cancer-associated skeletal muscle remodeling. However, this mechanism is likely less involved in early-stage disease when overall tumor burden is relatively low. Finally, behavioral changes associated with cancer diagnosis and treatment, including alterations in diet and physical activity, may further influence skeletal muscle through nutritional deficiency and physical deconditioning.^45^ Together, these observations suggest that skeletal muscle remodeling in non-metastatic breast cancer arises from the convergence of multiple stressors that disrupt metabolic homeostasis within the muscle microenvironment.

Because oxidative fibers play a central role in sustaining muscle contraction, their preferential atrophy may have important implications for the fatigue experienced by many breast cancer survivors. Although cancer-related fatigue is widely recognized as a multifactorial symptom involving central psychological and peripheral physiological contributors,^46^ skeletal muscle dysfunction may represent an important and underappreciated component of this syndrome. The present study demonstrates that structural and metabolic remodeling of skeletal muscle can occur at early disease stages, even in individuals without prior exposure to systemic therapy. Although the long-term persistence of these alterations in breast cancer remains unknown, studies in other contexts indicate that moderate muscle atrophy and weakness can persist for years after an inciting insult, even with targeted rehabilitation.^47^ Emerging evidence also suggests that skeletal muscle may retain long-term molecular pathology through persistent transcriptional and epigenetic reprogramming.^48^ Together, these observations raise the possibility that breast cancer-associated perturbations could produce durable structural and molecular changes in skeletal muscle that contribute to the prolonged fatigue frequently reported by breast cancer survivors. In addition to its long-term impact on survivorship, severe fatigue can also limit tolerance to systemic therapies and contribute to dose reductions or early treatment cessation.^12^ Despite the high prevalence of cancer-related fatigue and its substantial impact on quality of life, there are currently no approved targeted therapies for this condition. Defining the specific stressors and mechanisms that drive cancer-associated skeletal muscle remodeling, as well as the pathways linking these changes to fatigue and the mechanisms underlying their persistence into survivorship, will be essential for informing the development of effective therapeutic strategies.

Beyond the biological insights described here, this study establishes an integrated framework for profiling skeletal muscle using biopsies obtained during routine surgical procedures. By combining ST with high-throughput single-fiber morphometry, this approach enables simultaneous characterization of muscle fiber architecture and spatially resolved transcriptional alterations. To our knowledge, this represents the first application of ST to human skeletal muscle biopsies obtained from individuals with cancer. This strategy provides a scalable platform for interrogating skeletal muscle across diverse disease contexts, including cancer, aging, metabolic disease, genetic syndromes, and disuse.

Several limitations of the present study should be acknowledged. First, the cross-sectional design and modest cohort size limit our ability to establish causal relationships between breast cancer, treatment exposures, and the observed skeletal muscle remodeling. Although the inclusion of individuals with heterogeneous treatment histories reflects real-world clinical populations, exposure of some individuals to neoadjuvant chemotherapy prevents the precise determination of the isolated contributions of tumor biology and systemic therapy to the observed skeletal muscle pathology. In addition, molecular subtype and treatment exposure were highly correlated in this cohort, as all individuals with triple-negative breast cancer received neoadjuvant chemotherapy, necessitating larger studies to disentangle tumor subtype- and therapy-associated effects. The ST analyses were also performed on a limited number of tissue sections and thus should be interpreted primarily as a high-resolution spatial characterization of previously reported population-level effects,^14^ rather than independent individual-level inference. Mechanistically, while the ST and morphometric analyses provide insight into transcriptional and structural alterations within skeletal muscle, additional studies incorporating protein expression measurements, functional assessments, and larger individual-level analyses will be required to validate the biological consequences of these changes. Finally, because cancer-related fatigue is a multifactorial symptom influenced by the central nervous system, psychological, and systemic physiologic factors, the present study cannot determine the relative contribution of skeletal muscle remodeling to fatigue severity. Despite these limitations, the concordance of structural, transcriptional, and spatial findings presented here, together with prior bulk RNA-sequencing analyses of skeletal muscle in non-metastatic breast cancer,^14^ supports a model in which oxidative muscle fiber atrophy arises from both intrinsic mitochondrial dysfunction and extrinsic stromal signaling within the muscle microenvironment, potentially contributing to persistent fatigue in breast cancer survivorship.

## Methods

### Human Subjects and Pectoralis Major Biopsy Procurement

This study was conducted in accordance with the ethical standards for human subjects research outlined in the Declaration of Helsinki.^49^ The protocol was reviewed and approved by the West Virginia University Institutional Review Board on October 28, 2015 (IRB Protocol Number 1509848715R013). Informed consent for research participation, publication, and future use of research data was obtained from all participants prior to enrollment and tissue collection.

Individuals were eligible to participate if they were at least 18 years old, female, and willing to donate a small surgically resected pectoralis major muscle biopsy for research purposes. Participants with a history of neoadjuvant treatment, including radiation and/or systemic chemotherapy, were eligible to participate to best reflect real-world surgical cohorts. Individuals with a current or prior diagnosis of any cancer type other than breast or basal cell carcinoma of the skin were excluded.

To be eligible for the control arm, participants were required to be generally healthy with no current or prior diagnosis of breast cancer and to have a planned breast reduction, breast reconstruction, implant removal, or prophylactic mastectomy procedure during which the surgical approach would normally expose a portion of the pectoralis major muscle. To be eligible for the breast cancer arm, participants were required to have a confirmed diagnosis of breast cancer and a planned physician-initiated surgical procedure that would similarly expose a portion of the pectoralis major muscle.

Clinical variables, including surgical indication, breast cancer molecular subtype, age, surgical side, and neoadjuvant treatment history, were logged by a collaborating clinical research team and kept blinded from the basic science research team until completion of all primary analyses. Pectoralis major biopsies were obtained intraoperatively by the attending surgeon, promptly transported to the laboratory space on ice, and transferred to the basic science research team, who immediately prepared them for the downstream applications as detailed below.

### Tissue Processing, Embedding, and Serial Sectioning

All biopsies were processed immediately and frozen within 1 hour of excision. During the processing window, the tissue was wrapped in sterile saline-soaked gauze and maintained on ice to minimize degradation. Muscle biopsies prepared for histologic applications, such as immunofluorescent (IF) fiber-typing, spatial transcriptomics (ST), and Periodic acid-Schiff (PAS) staining, were trimmed as needed, aligned in cross-section under a dissecting microscope, placed in a small plastic mold, embedded in optimal cutting temperature (OCT) compound, and frozen in isopentane pre-cooled with liquid nitrogen. Blocks were then held on dry ice until airtight storage at -80 °C. All biopsies were analyzed within two years of collection.

For histologic applications, 10 µm sections were cut at standard cryosectioning temperatures for frozen skeletal muscle (approximately -18 to -22 °C) using Leica cryostat instruments. Sections were placed onto pre-cooled charged microscope slides (Fisherbrand™ Superfrost™ Plus) to prevent tissue thawing during placement. To maximize image registration quality, interleaved serial sections were prepared for alternating downstream applications (immunofluorescence [IF] and spatial transcriptomics [ST]; IF–ST–IF–ST–IF). To preserve RNA integrity and comply with ST best practices, freeze-thaw cycles were intentionally avoided. Slides were stored in airtight slide mailers at -80 °C for up to 4 weeks to preserve tissue integrity prior to downstream processing.

### Immunofluorescent Fiber-Typing and Imaging

IF skeletal muscle fiber-typing was performed based on an established protocol described by Murach et al.,^50^ with specific modifications detailed below. Cryosections prepared as described above were used for IF staining. Sections were thawed and air-dried at room temperature (RT) for at least 1 h prior to staining. Sections exhibiting folds or tears were excluded prior to staining as part of routine histologic quality control.

A primary antibody master mix was prepared in phosphate-buffered saline (PBS) supplemented with 1% bovine serum albumin (BSA; w/v) according to the antibody concentrations outlined in Supplementary Table 3. Consistent with the previously described protocol, the primary antibody master mix was applied directly to tissue sections without fixation or blocking and incubated at 4 °C in a humidified slide box overnight. The primary antibody master mix was removed by pipetting, and slides were washed 3 times for 5 min each in PBS at RT. Sections were then briefly air dried, and a secondary antibody master mix was prepared in PBS containing 1% BSA (w/v) according to the antibody concentrations outlined in Supplementary Table 4. The secondary antibody master mix was applied directly to tissue sections and incubated for 1 h at RT in a humidified slide box protected from light. The secondary antibody master mix was then removed, and sections were washed using the same procedure described above. Sections were then mounted and coverslipped using VECTASHIELD Vibrance Antifade Mounting Medium (Vector Laboratories; H-1700-10; RRID:AB_2336789) and cured overnight at RT.

Whole tissue sections were imaged using an Olympus VS120 widefield slide-scanning fluorescence microscope. Imaging was performed in a single optical plane using a 20x, 0.75-numerical-aperture (NA) air objective. Tiled images were acquired using an Olympus XM10 monochrome CCD camera and stitched using the manufacturer’s software. The resulting square pixel size for all images was 0.3239 µm per pixel in both dimensions. Four fluorescence channels were acquired sequentially using empirically determined fixed exposure times for each staining batch. Across staining batches, exposure times were adjusted to account for batch-to-batch staining variation and optimally expose all four fluorescence channels. Therefore, relative fluorescence intensities were not compared across staining batches. Uncompressed images for downstream analysis were exported as four-channel, 16-bit grayscale TIFF files without any intensity transformation.

### Serial Section Image Alignment

ST was performed on hematoxylin and eosin (H&E) stained sections, with IF fiber-typing performed on two serially adjacent sections. To enable meaningful cross-modal image alignment, sections were selected for ST based on having consistent morphology with at least one adjacent IF section. Given the longitudinal continuity of skeletal muscle fibers, the same muscle fibers were present in both sections.

Adjacent whole section H&E and IF images were acquired using the same microscope and imaging configurations described above in brightfield and fluorescence modes, respectively. Images were aligned using the 10x Genomics Loupe Browser Visium Image Alignment tool (v8.1.2; RRID:SCR_018555). Although this tool is not formally validated for use with serial sections, it was used here to establish approximate spatial correspondence between modalities for downstream visualization.

Image alignment was exclusively used to facilitate cross-modal visualization of ST transcriptional spots and IF-typed skeletal muscle fibers. ST cluster assignment and spot annotation were based solely on ST-derived gene expression data and established muscle fiber-type marker genes and did not depend on the relative alignment of ST spots to adjacent IF sections. Concordance between transcriptionally defined ST clusters and adjacent IF-typed skeletal muscle fibers was assessed qualitatively, revealing clear agreement between modalities (Fig. 2H). Given the use of adjacent sections, analyses were intentionally structured to minimize dependence on precise spatial image alignment, and no conclusions rely on the relative positioning of ST spots to adjacent IF-typed skeletal muscle fibers.

### Skeletal Muscle Fiber Segmentation

To accurately segment cross-sectional skeletal muscle fibers from whole-tissue IF images, we developed a custom Python-based image-processing pipeline implemented with Dask (RRID:SCR_026688) to enable parallel processing and limit peak RAM requirements for large stitched images. The segmentation logic was conceptually inspired by Wen et al.,^51^ with modifications to enable processing of large stitched images and automated muscle fiber-typing. The authors’ original implementation is available on GitHub.^52^ Our pipeline accepts multichannel BigTIFF files (.btf) containing high-resolution IF images of skeletal muscle. Each file stores multiple two-dimensional fluorescence channels acquired from the same tissue section with spatial co-registration. For this project, the channels included a laminin stain to detect muscle fiber boundaries and three fiber-type-specific myosin heavy chain stains to identify muscle fiber types.

Muscle fiber boundary detection and seed generation were accomplished using a combination of image processing and morphologic operations defined relative to the image pixel scale. Steps included laminin channel normalization and percentile-based contrast enhancement, tissue mask generation, Frangi filter^53^ edge detection, and muscle fiber seeding via thresholding and conversion to a binary image map. Muscle fiber seeds were then refined by filling internal holes and removing islands that were too small to be candidate fibers. At this stage, segmentation seeds represented candidate muscle fiber interiors and were used for machine-learning-based classification and morphologic refinement, as described below.

### Machine Learning-Based Muscle Fiber Classification and Segmentation Refinement

For each candidate fiber, 9 morphometric parameters describing fiber geometry and 26 channel-intensity features capturing staining characteristics across laminin and myosin heavy chain channels were calculated. Two histogram-based gradient boosting classifiers implemented in scikit-learn^54^ (RRID:SCR_002577) were trained on these parameters using 4,800 manually annotated candidate muscle fibers. Histogram-based gradient boosting classifiers were selected for their ability to model nonlinear feature interactions without scaling, robustness to heterogeneous feature distributions, and computational efficiency on moderately sized datasets.

The first model utilized all 35 features to classify segmentation masks into one of six classes: high-quality cross-sectional muscle fiber, medium-quality cross-sectional muscle fiber, low-quality muscle fiber, longitudinal muscle fiber, merged muscle fibers, or other structure. These classes are hereafter abbreviated as high-quality (HQ), medium-quality (MQ), low-quality (LQ), longitudinal (Long), merged (Merge), and other structure (Other), respectively. To maximize precision for the high-quality class and preferentially route borderline segmentations to morphologic refinement, a conservative decision threshold (𝜏=0.815) was applied to this class during the initial classification stages.

At each stage of classification, segmentations labeled as high-quality were locked and saved for morphometric analysis; segmentations classified as low-quality, longitudinal, or other structures were excluded; segmentations classified as medium-quality or merged were subjected to iterative morphologic refinement.

Refinement consisted of a series of morphologic operations, including binary opening and closing for medium-quality segmentations and watershed-based merge splitting for merged segmentations. Following each refinement step, the same classifier was used to re-classify the modified candidates, and segmentations were sorted according to the decision logic described above.

Refinement iterations continued until either fewer than 10% of candidate segmentations were converted to the high-quality class on 2 consecutive iterations or a maximum of 10 refinement iterations had been completed. Once morphologic refinement was complete, the conservative decision threshold applied to the high-quality class was removed, and final classifications were assigned based on the model’s maximum predicted probability. Any remaining medium-quality or merged segmentations were excluded from morphometric analysis.

A second model was trained on manually annotated high-quality fibers and utilized 24 channel intensity statistics derived from myosin heavy chain IF channels to classify segmentation masks as one of three fiber types: type I (red), type IIa (green), or type IIx (blue). This second model was only applied to segmentations classified as high-quality by the first model after morphologic refinement was complete. Morphometric parameters were excluded from this model to avoid biasing fiber-type classification by fiber geometry.

### Machine Learning Model Validation and Performance

Classifier performance was evaluated independently from the morphologic refinement procedure to assess intrinsic model performance. Performance metrics for both models, including precision-recall (PR) curves, receiver operating characteristic (ROC) curves, confusion matrices, and summary performance statistics, are presented in Supplementary Fig. 1. The models were evaluated on 1,200 manually annotated segmentation masks that were not included in the training dataset. The first classifier demonstrated high precision for the high-quality class (precision=0.85) while using the conservative operating threshold during the initial classification iterations. In addition, model outputs were visually inspected across multiple independent sections to confirm classification accuracy and identify potential segmentation artifacts. Representative examples illustrating classifier outputs before and after morphologic refinement are shown in Supplementary Fig. 1H. Importantly, classifier performance was comparable between non-cancer and breast cancer muscle sections. This validation confirmed that model-derived segmentations were sufficiently accurate to support downstream morphometric analyses.

### Single-Fiber Skeletal Muscle Morphometric Analysis

Morphometric analyses were performed on segmentation masks that were classified as high-quality after morphologic refinement. Cross-sectional area and minor axis length were extracted for each fiber and compared at the single-fiber level between the non-cancer and breast cancer cohorts. Additional analyses were performed to evaluate potential associations between morphometric parameters and clinical variables, including AJCC clinical anatomic stage, breast cancer molecular subtype, neoadjuvant chemotherapy exposure, age, and BMI. An exploratory analysis was also conducted to evaluate correlations between muscle fiber morphometric parameters and participant-reported fatigue (FACIT-F fatigue subscale scores). To assess the robustness of the single-fiber analysis, a complementary individual-level analysis was performed by excluding sections that contained fewer than 1,000 high-quality fibers, averaging morphometric variables per individual, and comparing group-level differences.

### Spatial Transcriptomics Sample Selection, Library Preparation, and Sequencing

To select sections for ST, tissue blocks were first evaluated for RNA integrity by collecting ten 10 µm sections, trimming excess OCT, and extracting total RNA using the Qiagen RNeasy Fibrous Tissue Mini Kit according to the manufacturer’s protocol. Total RNA was analyzed on an Agilent 4150 TapeStation (RRID:SCR_019393) to determine each block’s RNA integrity number (RIN). All profiled samples exceeded the 10x Genomics recommended threshold of 4.0 for probe-based assays, with RIN scores ranging from 5.6 to 7.4. Sections from blocks with acceptable RNA integrity were then evaluated for morphologic consistency with adjacent IF muscle sections to guide the final selection of sections for ST profiling.

A total of 4 ST libraries were generated using the probe-based 10x Genomics Visium CytAssist Spatial Gene Expression kit for fresh frozen tissues (v2; 6.5 x 6.5 mm capture area; RRID:SCR_024570).^55^ Tissue sections prepared as described above were processed for probe hybridization, ligation, extension, pre-amplification, and library construction according to the manufacturer’s protocol. CytAssist procedures were performed by the West Virginia University Flow Cytometry & Single Cell Core Facility, while the final library construction was performed by the West Virginia University Genomics Core Facility. Sequencing was performed by Admera Health LLC (South Plainfield, NJ, USA) on an Illumina NovaSeq X Plus platform using a 2 x 150 base-pair read configuration, exceeding the minimum read lengths specified by the Visium read configuration. Libraries were sequenced to a target average depth of >50,000 reads per tissue-covered spot, with actual sequencing depth ranging from 51,547 to 93,601 reads per tissue-covered spot.

### Spatial Transcriptomics Preprocessing and Quality Control

Raw sequencing data were initially processed using the 10x Genomics Space Ranger pipeline (version 3.1.2; RRID:SCR_025848), which performs tissue detection, image registration, spatial barcode demultiplexing, read alignment to the reference probe set, UMI-based transcript counting, and generation of spot-level gene expression matrices. Inputs included the 10x Genomics Visium Human Transcriptome Probe Set (version 2.0) and image alignment JSON files generated by the 10x Genomics Loupe Browser Visium Image Alignment tool that encoded the spatial alignment between the Visium ST spots and the adjacent IF section.

Downstream preprocessing and quality control were performed in Python (version 3.11.11; RRID:SCR_008394) using Scanpy (version 1.11.4; RRID:SCR_018139)^56^ and Squidpy (version 1.6.2; RRID:SCR_026157)^57^. Space Ranger output matrices were imported using the read.visium function in Squidpy. Libraries were aggregated into a single AnnData object and filtered to remove spots with fewer than 10,000 UMIs, fewer than 2,000 detected genes, or more than 10% mitochondrial gene expression. Filtering thresholds were selected to remove low-quality and background spots. Filtered data were visually inspected prior to downstream analyses to ensure spatial coherence between Visium spots and tissue-covered regions, as seen in Supplementary Figures 4-5. Counts were then normalized to a target sum of 10,000 counts per spot, excluding highly expressed genes, and subsequently log-transformed.

### Spatial Transcriptomics Batch Correction, Clustering, and Annotation

Prior to clustering, residual technical variance was further reduced using the regress_out function in Scanpy over known technical covariates, including UMI counts, number of detected genes, and percent mitochondrial gene expression per spot. The corrected expression values were then centered and scaled to unit variance. Highly variable genes (HVG) were subsequently identified across the aggregated dataset using the Seurat flavor^58^ of the highly_variable_genes function in Scanpy, with library identity treated as a batch variable to ensure library-aware feature selection. Downstream clustering was restricted to the top 2,000 highly variable genes.

Principal component analysis (PCA) was performed on the HVG-restricted, regressed, and scaled expression values described above. To mitigate library-level batch effects while preserving shared biological information, Harmony^59^ integration was applied to the first 50 principal components using library identity as the grouping variable. A k-nearest neighbor graph (n = 15) was constructed from the Harmony-corrected PCA space. Two-dimensional embeddings were generated using uniform manifold approximation and projection (UMAP),^60^ and clusters were identified using the Leiden algorithm^61^ at a resolution of 0.4. Cluster quality was assessed by visualizing overlays of known technical covariates on the UMAP embedding (Supplementary Fig. 6).

While clustering was performed on the HVG-restricted, regressed, and scaled expression layer, differentially expressed marker genes were identified from the log-normalized expression layer using the rank_genes_groups function with a t-test and a Benjamini-Hochberg multiple-testing correction.^62^ Clusters were annotated based on differential expression of canonical cell-type marker genes. Myofiber subtypes were further classified using previously reported transcriptional markers for type I and type IIa skeletal muscle fibers,^63^ with visual cross-validation against background IF images where appropriate.

### Spatial Transcriptomics Differential Expression and Gene-Set Enrichment Analyses

Differential expression analysis between the non-cancer and breast cancer conditions was performed separately for each annotated spot-type using log-normalized expression values. Statistical testing was performed using a two-sided t-test with Benjamini–Hochberg correction for multiple comparisons.

For gene-set enrichment analysis (GSEA), gene rankings were derived from signed t-statistics generated during differential expression testing. The resulting rank lists were used as inputs to blitzGSEA (version 1.3.40),^64^ and enrichment analyses were performed using the KEGG 2021 Human gene set database.^65^ To assess gene-set enrichment with spot-level resolution, per-spot gene signature scores were computed using UCell (version 0.5.0; RRID:SCR_027109)^23,24^ and projected onto tissue sections to visualize spatial patterns in pathway activity.

### Spatial Ligand-Receptor Proximity-Enrichment Analysis

To evaluate spatially localized ligand-receptor proximity enrichment, a spatial neighbors graph was constructed using the spatial_neighbors function in Squidpy, with two concentric rings of neighboring spots. Candidate ligand-receptor interactions were assessed between adipocyte-annotated sender spots expressing predefined ligands and myofiber-annotated (Type I, Type IIa, and MB) receiver spots expressing cognate receptors within each library. For each ligand-receptor pair, a spatial coupling score was computed as the mean ligand-receptor log-normalized expression product across neighboring sender-receiver spot pairs. To account for differences in overall ligand and receptor abundance, coupling scores were normalized by the product of the mean ligand expression in sender spots and the mean receptor expression in receiver spots within the same library. Normalized scores were log2-transformed and averaged within each condition for visualization.

### PAS Staining and Blinded PAS Reactivity Assessment

PAS staining was performed by the West Virginia University Pathology Research Histology Core Facility on 10 µm sections obtained from the same blocks used for ST profiling and IF muscle fiber-typing. Tissue sections were stained using a standard clinical histopathology protocol for PAS detection of glycogen and other carbohydrate-rich structures. Whole slide images were obtained by brightfield microscopy using a 20x, 0.75-numerical-aperture (NA) air objective on an Olympus VS120 slide-scanning microscope. Tiled images were acquired using an Olympus XM10 monochrome CCD camera and stitched using the manufacturer’s software. Representative regions of interest were selected by an investigator blinded to sample identity and qualitatively assessed for PAS reactivity. Representative images are included in Supplementary Figure 3.

### Statistical Analysis

Statistical analyses were performed in Python (version 3.11.11) using Scanpy (version 1.11.4), SciPy (version 1.16.1), and Statsmodels (version 0.14.5; RRID:SCR_016074).^66^ Unless otherwise stated, statistical testing was performed at the fiber or ST spot level. Data distributions were visualized using Seaborn (version 0.13.2; RRID:SCR_018132)^67^ split violin plots with embedded box-and-whisker plots and individual participant means overlaid. Two-sided statistical tests were used for all comparisons. For single fiber morphometry comparisons between two groups, Welch’s t-tests were used to account for unequal variances. For ST comparisons between two groups, Mann-Whitney U tests were used to account for non-normality. For analyses with multiple comparisons, P values were adjusted using a Benjamini-Hochberg false discovery rate (FDR) correction. Statistical significance was defined as adjusted P < 0.05. Exact statistical tests, units of comparison, and adjusted P value thresholds are reported in the corresponding figure legends.

## Data and Code Availability

Unprocessed ST FASTQ files and Space Ranger outputs, including raw and filtered feature-barcode matrices, have been deposited in the National Center for Biotechnology Information (NCBI) Sequence Read Archive (SRA) and Gene Expression Omnibus (GEO) repositories, respectively, under the BioProject accession number PRJNA1458763.

Associated spatial metadata, including images, spatial coordinates, scale factors, and alignment JSON files used as Space Ranger inputs are available on Zenodo (10.5281/zenodo.19825818). Spatial transcriptomics datasets are additionally provided as AnnData (.h5ad) objects on Zenodo (10.5281/zenodo.19825818), including both minimally processed versions generated directly from the Space Ranger outputs and aggregated, fully processed versions with clustering and annotation.

Multi-channel IF images used for single-fiber morphometry and the corresponding morphometric datasets are also available on Zenodo (10.5281/zenodo.19858383).

Code used for analysis and figure generation, including trained machine-learning models for single-fiber segmentation and muscle fiber-typing, is available on GitHub: https://github.com/PistilliLab/Single-fiber-morphometry-and-spatial-transcriptomics-of-human-breast-cancer-muscle. A version of this repository archived at the time of publication will also be made available on Zenodo (TBD). Generative AI tools (ChatGPT, OpenAI) were used to assist with the development and testing of custom Python scripts. All generated code and outputs were reviewed and validated by the authors.

To protect participant privacy, individual-level continuous clinical variables including participant age, BMI, and FACIT-F scores will not be made publicly available. Portions of the analysis code requiring restricted clinical data are annotated accordingly.

All data deposited in the above repositories will be made publicly accessible at the time of final publication. All other data will be made available upon reasonable written request to the corresponding author.

## Acknowledgements

The authors would like to thank all of the study participants for their contributions to this research. The following sources provided funding for this work: National Institute of Arthritis and Musculoskeletal and Skin Diseases (NIAMS; R01AR079445; Pistilli) and the West Virginia University (WVU) Cancer Institute. We acknowledge the WVU Flow Cytometry & Single Cell Core Facility (NIGMS; P30GM121322, P20GM103434), the WVU Genomics Core Facility, the WVU Microscope Imaging Facility (NIH; P20GM121322, P20GM144230, P20GM103434), and the WVU Pathology Research Histology Core Facility for assistance with data acquisition. We thank Nick Hanna, Pratikshya Shrestha, Joshua Gibson, Gabriel Sankey, and Colin McBee for contributing muscle fiber annotations used to train the ML models. We also thank Emily Hawkins, Kaitlyn Hager, and the rest of the WVU Cancer Institute Clinical Research Unit for identifying, enrolling, and tracking all of the participants in this study.

## Author Contributions

Conceptualization: ADM, HEW, HHJ, JFP, and EEP; Methodology: ADM; Software: ADM and SAC; Investigation: ADM, SAC, ALB, IAO, MAW, EEP; Resources: MAW; Data curation: ADM, ALB, and IAO; Formal analysis: ADM; Visualization: ADM; Writing – original draft: ADM; Writing – review & editing: all authors; Supervision: HHJ, JFP, and EEP; Funding acquisition: EEP.

