## Supplemental Information for "Single-fiber morphometry and spatial transcriptomics reveal selective oxidative muscle fiber atrophy in non-metastatic breast cancer"

### Supplementary Table 1 – Individual participant clinical metadata.

| Manuscript ID | Study arm | AJCC clinical anatomic stage | Molecular subtype | NACT regimen |
|---|---|---|---|---|
| BC1 | Breast cancer | I | Luminal | None |
| BC2 | Breast cancer | I | Triple positive | None |
| BC3 | Breast cancer | II | Triple positive | TCHP |
| BC4 | Breast cancer | III | HER2-positive | TCHP |
| BC5 | Breast cancer | I | Luminal | None |
| BC6 | Breast cancer | II | HER2-positive | None |
| BC7 | Breast cancer | III | Triple negative | KEYNOTE-522 |
| BC8 | Breast cancer | II | Triple positive | THP |
| BC9 | Breast cancer | II | Triple negative | KEYNOTE-522 |
| BC10 | Breast cancer | II | Triple positive | TCHP |
| BC11 | Breast cancer | III | HER2-positive | THP |
| BC12 | Breast cancer | II | Luminal | None |
| BC13 | Breast cancer | III | Triple negative | KEYNOTE-522 |
| BC14 | Breast cancer | I | Luminal | None |
| BC15 | Breast cancer | III | HER2-positive | TCHP |
| BC16 | Breast cancer | II | HER2-positive | TCHP |
| NC1 | Non-cancer | — | — | — |
| NC2 | Non-cancer | — | — | — |
| NC3 | Non-cancer | — | — | — |
| NC4 | Non-cancer | — | — | — |

AJCC, American Joint Committee on Cancer; NACT, neoadjuvant chemotherapy; Luminal (hormone receptor-positive [HR+]/human epidermal growth factor receptor 2-negative [HER2–]); HER2-positive (HR–/HER2+); triple-positive (HR+/HER2+); triple-negative (HR–/HER2–). TCHP, docetaxel/carboplatin/trastuzumab/pertuzumab; THP, docetaxel/trastuzumab/pertuzumab; KEYNOTE-522, pembrolizumab/paclitaxel/carboplatin followed by pembrolizumab/doxorubicin/cyclophosphamide.

**Supplementary Table 2 – Sample utilization by assay.**

| Manuscript ID | Used for ST | Used for IF | Used for PAS |
|---|---|---|---|
| BC1 | Yes | Yes | Yes |
| BC2 | Yes | Yes | Yes |
| BC3 | No | Yes | Yes |
| BC4 | No | Yes | Yes |
| BC5 | No | Yes | Yes |
| BC6 | No | Yes | Yes |
| BC7 | No | Yes | Yes |
| BC8 | No | Yes | Yes |
| BC9 | No | Yes | Yes |
| BC10 | No | Yes | Yes |
| BC11 | No | Yes | Yes |
| BC12 | No | Yes | Yes |
| BC13 | No | Yes | Yes |
| BC14 | No | Yes | Yes |
| BC15 | No | Yes | Yes |
| BC16 | No | Yes | Yes |
| NC1 | Yes | Yes | Yes |
| NC2 | Yes | Yes | Yes |
| NC3 | No | Yes | Yes |
| NC4 | No | Yes | Yes |

ST, spatial transcriptomics; IF, immunofluorescent muscle fiber-typing; PAS, periodic acid-Schiff staining.

**Supplementary Table 3 – Primary Antibodies.**

| Primary Antibodies | | | | | | | | | |
|---|---|---|---|---|---|---|---|---|---|
| **Target** | **Gene Symbol** | **Clone** | **Host Species** | **Isotype** | **Vendor** | **Catalog Number** | **RRID** | **Final Staining Concentration (µg/mL)** | **Incubation** |
| **MyHC I** | MYH7 | BA-D5 | Mouse | IgG2b | DSHB | BA-D5 | RRID: AB_2235587 | 3.05 | O/N, 4 ºC |
| **MyHC IIA** | MYH2 | SC-71 | Mouse | IgG1 | DSHB | SC-71 | RRID: AB_2147165 | 0.85 | O/N, 4 ºC |
| **MyHC IIX** | MYH1 | 6H1 | Mouse | IgM | DSHB | 6H1 | RRID: AB_1157897 | 10.5 | O/N, 4 ºC |
| **Laminin*** | LAMA1 LAMB1 LAMC1 | Poly-clonal | Rabbit | IgG | Invitrogen | PA1-16730 | RRID: AB_2133633 | 10.4 | O/N, 4 ºC |

* - Laminin antibody (PA1-16730) was raised against laminin 111 isolated from mouse Engelbreth-Holm-Swarm (EHS) sarcoma cells and is not isoform-specific; DSHB, Developmental Studies Hybridoma Bank; O/N, overnight.

### Supplementary Table 4 – Secondary Antibodies.

| Secondary Antibodies | | | | | | | |
|---|---|---|---|---|---|---|---|
| **Specificity** | **Fluorophore** | **Host Species** | **Vendor** | **Catalog Number** | **RRID** | **Final Staining Concentration (µg/mL)** | **Incubation** |
| **Mouse IgG2b** | Alexa Fluor 647 | Goat | Invitrogen | A21242 | RRID:AB_2535811 | 10 | 1 h, RT |
| **Mouse IgG1** | Alexa Fluor 488 | Goat | Invitrogen | A21121 | RRID:AB_2535764 | 10 | 1 h, RT |
| **Mouse IgM** | Alexa Fluor 555 | Goat | Invitrogen | A21426 | RRID:AB_2535847 | 10 | 1 h, RT |
| **Rabbit IgG** | Alexa Fluor 405 | Goat | Invitrogen | A48254 | RRID:AB_2890548 | 20 | 1 h, RT |

RT, room temperature.

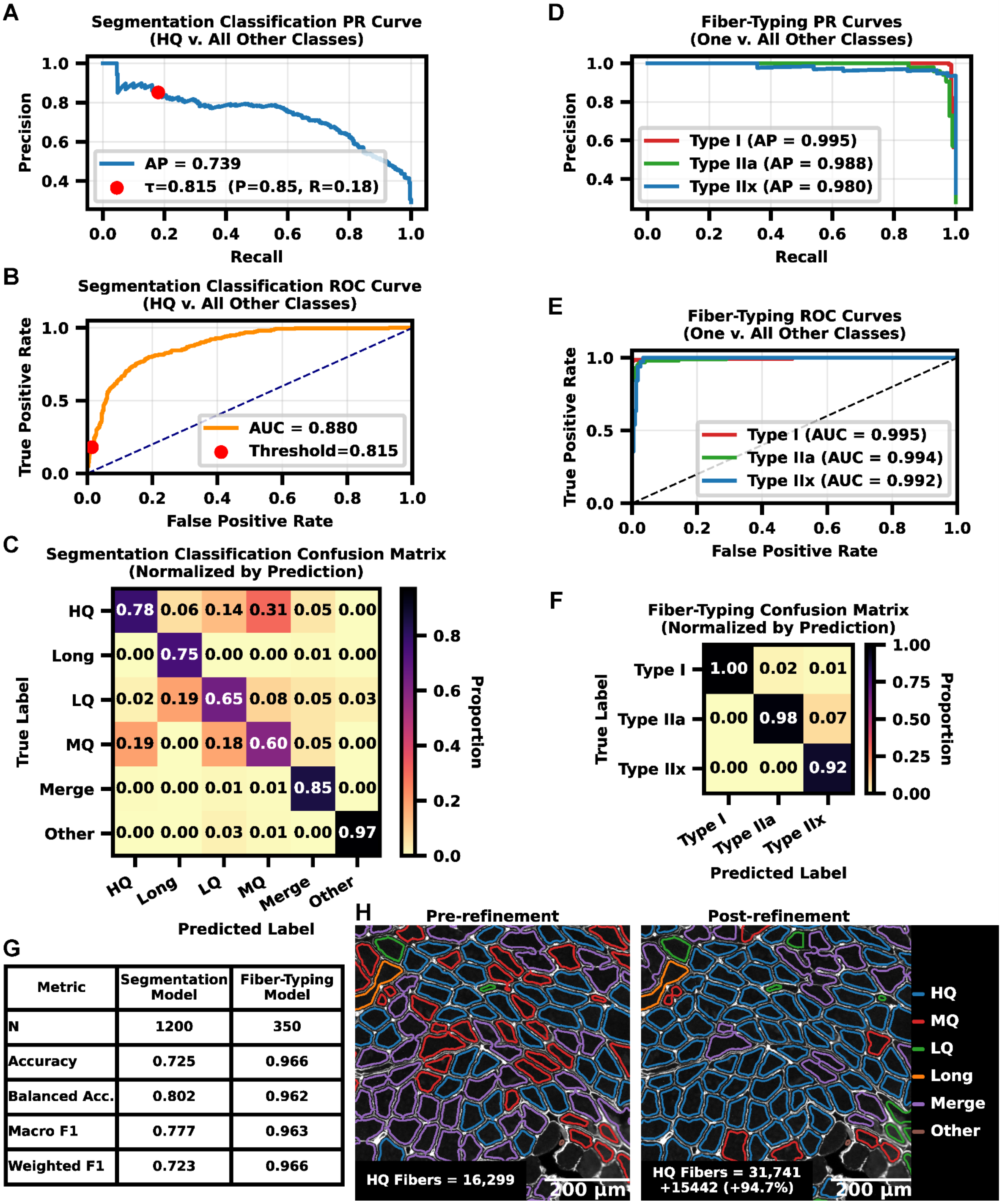


| Metric | Segmentation Model | Fiber-Typing Model |
|---|---|---|
| N | 1200 | 350 |
| Accuracy | 0.725 | 0.966 |
| Balanced Acc. | 0.802 | 0.962 |
| Macro F1 | 0.777 | 0.963 |
| Weighted F1 | 0.723 | 0.966 |

**Supplementary Figure 1 – Machine learning single fiber morphometry models effectively identify high-quality cross-sectional muscle fibers and assign muscle fiber type.**

**A** Precision-recall (PR) curve for the muscle fiber segmentation model. AP, average precision across all decision thresholds; P, precision; R, recall. **B** Receiver operating characteristic (ROC) curve for the muscle fiber segmentation model. **A-B** Curves are for the high-quality cross-sectional muscle fiber (HQ) class versus all other classes. Dots represent the conservative decision threshold ($\tau$=0.815) used during iterative morphologic refinement. **C** Confusion matrix for the muscle fiber segmentation model. **D** Precision-recall (PR) curves for the muscle fiber-typing model. AP, average precision across all decision thresholds. **E** Receiver operating characteristic (ROC) curves for the muscle fiber-typing model. **D-E** Curves are for the indicated class versus all other classes. **F** Confusion matrix for the muscle fiber-typing model. **G** Summary performance metrics for the muscle fiber segmentation and muscle fiber-typing models. N, number of candidate fibers in the validation test dataset; Accuracy, overall proportion correct; Balanced acc., mean recall across all classes; Macro F1, unweighted mean of class-specific F1 scores (harmonic mean of precision and recall for each class); Weighted F1, class-weighted average of class-specific F1 scores (corrects for underlying class imbalance). **H** Representative segmentation classifications before (left) and after (right) iterative morphologic refinement. Classifications were assigned by maximum predicted label probability. **A-G** Performance metrics calculated using a random 25% holdout set of manual annotations not included in the training dataset. **C,F** Confusion matrices were generated using classifications assigned by maximum predicted label probability and are normalized by predicted label (columns sum to 1.0). **C,H** HQ=high-quality cross-sectional muscle fiber; MQ=medium-quality cross-sectional muscle fiber; LQ=low-quality muscle fiber; Long=longitudinal muscle fiber; Merge=merged muscle fiber; Other=other structure.

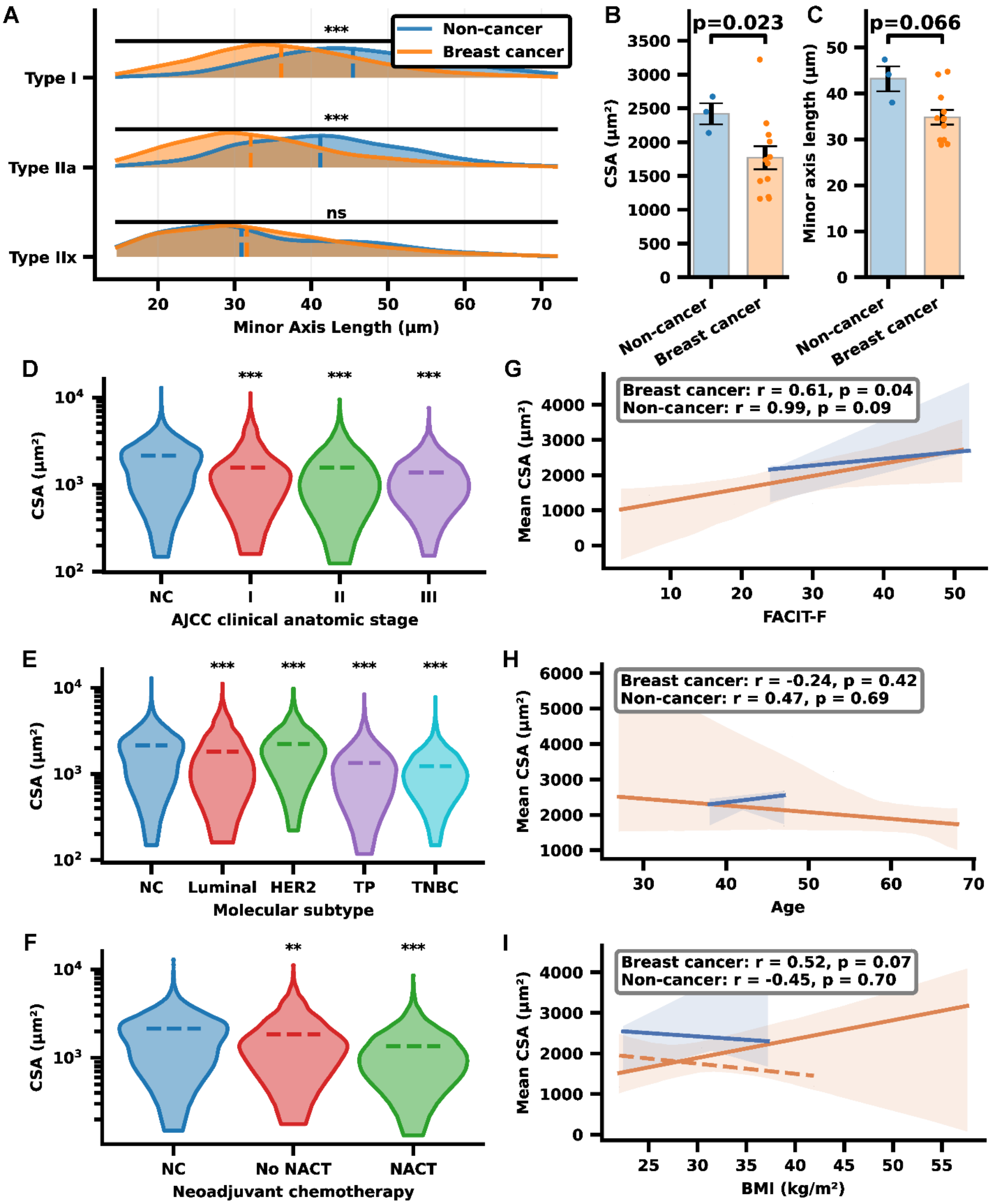

A
Non-cancer
Breast cancer
***
***
ns
Type I
Type IIa
Type IIx
Minor Axis Length (μm)
B
p=0.023
CSA (μm²)
C
p=0.066
Minor axis length (μm)
Non-cancer
Breast cancer
D
CSA (μm²)
NC
I
II
III
AJCC clinical anatomic stage
G
Breast cancer: r = 0.61, p = 0.04
Non-cancer: r = 0.99, p = 0.09
Mean CSA (μm²)
FACIT-F
E
NC
Luminal
HER2
TP
TNBC
Molecular subtype
H
Breast cancer: r = -0.24, p = 0.42
Non-cancer: r = 0.47, p = 0.69
Age
F
NC
No NACT
NACT
Neoadjuvant chemotherapy
I
Breast cancer: r = 0.52, p = 0.07
Non-cancer: r = -0.45, p = 0.70
BMI (kg/m²)

**Supplementary Figure 2 – Muscle fiber cross-sectional area correlates with participant-reported fatigue and other clinical variables.**

**A** Stratification of muscle fiber minor axis length by fiber type. Dashed lines represent medians. **B-C** Individual-level comparisons of cross-sectional area (CSA; **B**) and minor axis length (**C**) after excluding one sample that was an outlier with respect to BMI. Sections with >1000 high-quality fibers were used to compute per-individual means. n = individual study participants. Dots represent individual participant means. Error bars represent means ± standard error of the mean. **D-F** Comparison of muscle fiber cross-sectional area stratified by American Joint Committee on Cancer (AJCC) clinical anatomic stage (**D**), breast cancer molecular subtype (**E**), and neoadjuvant chemotherapy (NACT) exposure (**F**). All comparisons were pairwise between the experimental group (marked with asterisks) and the non-cancer (NC) control group. n = single muscle fibers. Dashed lines represent medians. CSA, muscle fiber cross-sectional area; NC, non-cancer; I, AJCC clinical anatomic stage 1; II, AJCC clinical anatomic stage 2; III, AJCC clinical anatomic stage 3; Luminal (hormone receptor-positive [HR+]/human epidermal growth factor receptor 2-negative [HER2–]); HER2 (HR–/HER2+); TP (HR+/HER2+); TNBC (HR–/HER2–). **G-I** Scatter plots depicting correlations between individual mean muscle fiber cross-sectional area (y-axis) and participant Functional Assessment of Chronic Illness Therapy–Fatigue (FACIT-F) subscale scores (**G**), participant age (**H**), and body mass index (BMI; **I**). Pearson correlation coefficients (r) and corresponding two-sided p values (null hypothesis = no linear association) are shown. n = individual study participants. Solid lines represent cohort linear regression lines. Shaded regions represent the 95% confidence interval. **I** The dashed line represents the regression line for the breast cancer cohort after excluding the sample with an associated BMI that exceeded the upper Tukey outlier limit. ($r = -0.26$; $p = 0.41$). **A-F** Comparisons between cohorts were performed using a two-tailed Welch's t-test. *$P < 0.05$; **$P < 0.01$; ***$P < 0.001$.

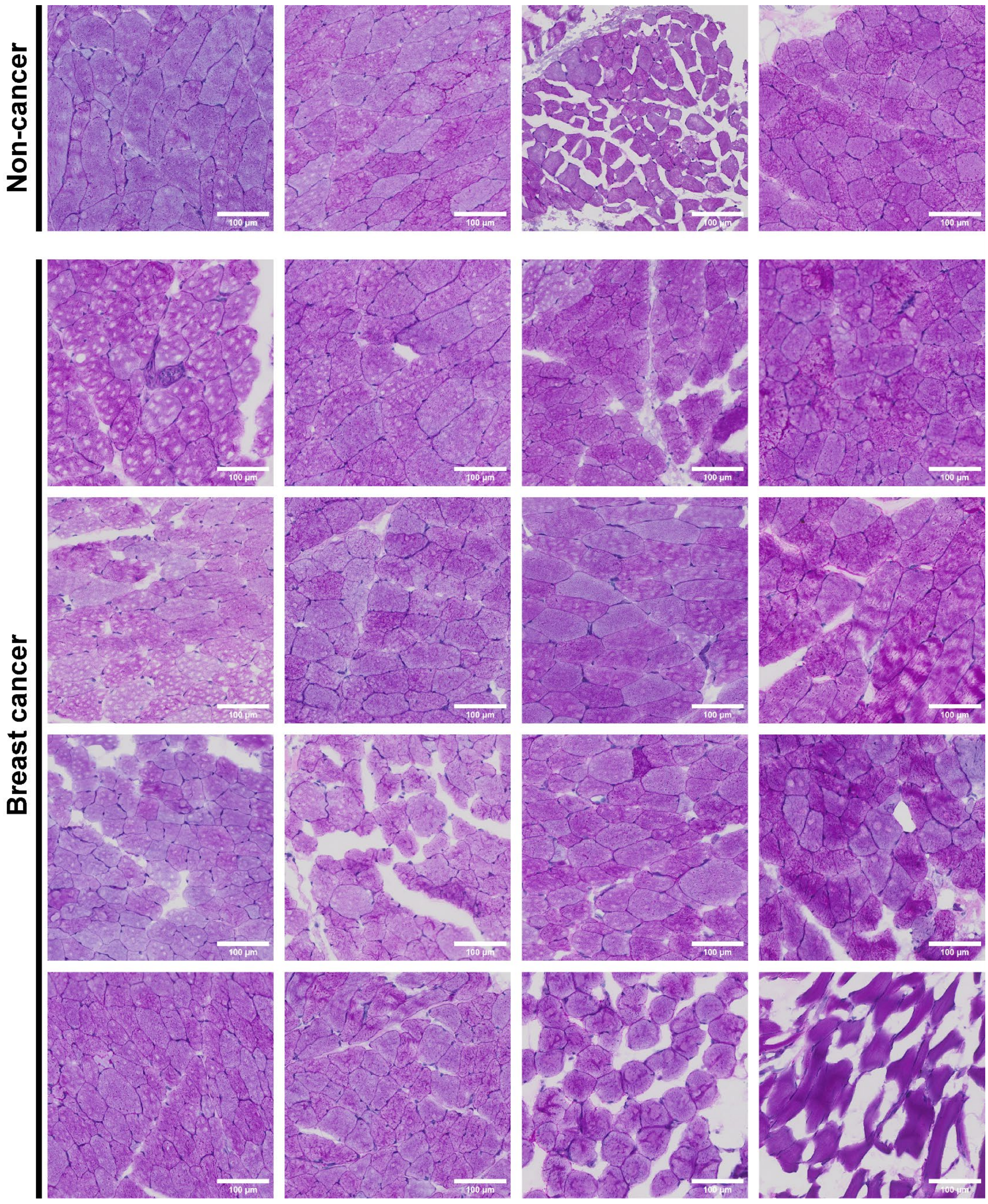
Non-cancer
Breast cancer
100 µm

**Supplementary Figure 3 – Representative images of periodic acid-Schiff (PAS) stained pectoralis major muscle biopsy sections.**

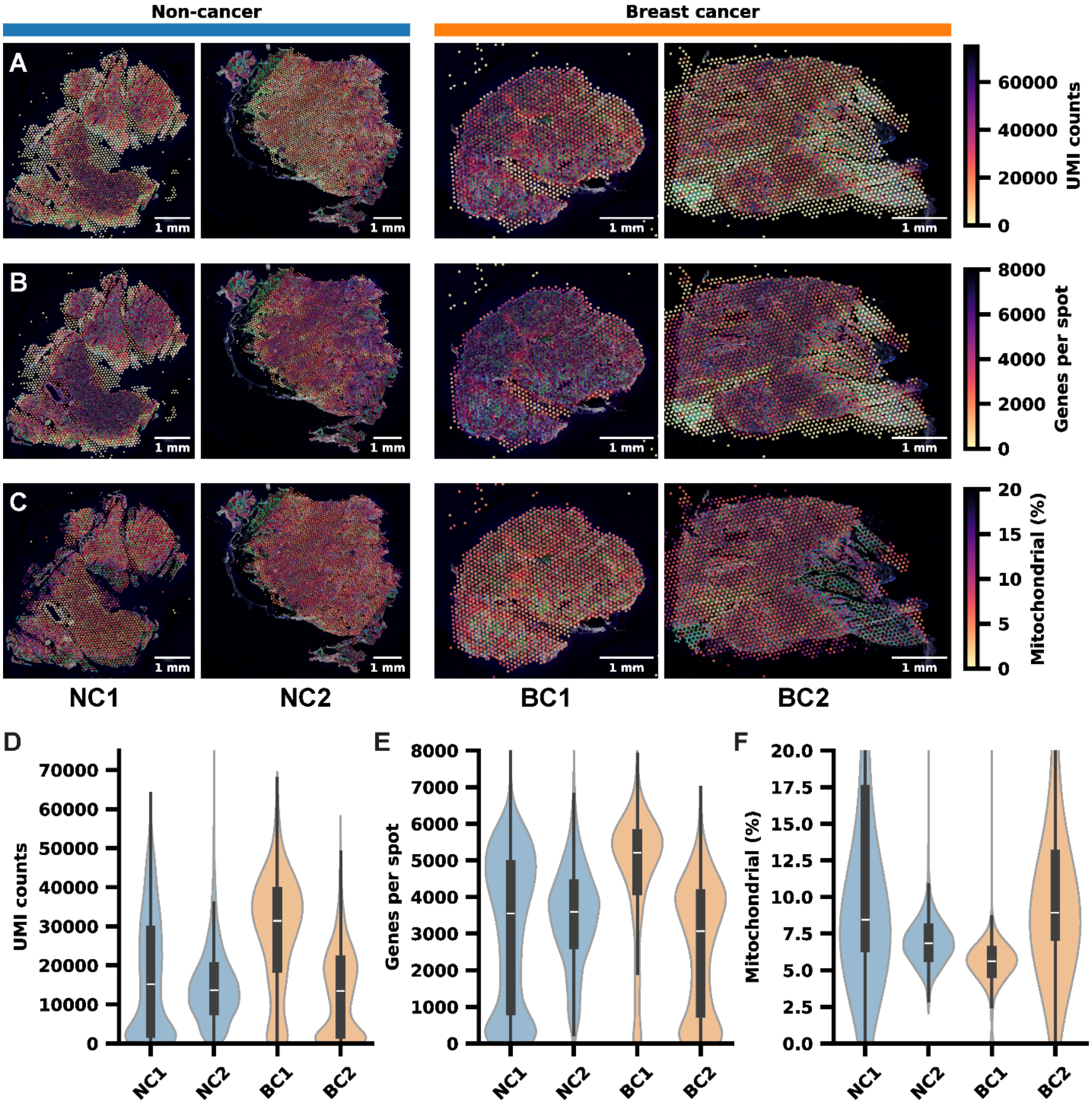
Non-cancer
Breast cancer
A
B
C
1 mm
UMI counts
0
20000
40000
60000
Genes per spot
0
2000
4000
6000
8000
Mitochondrial (%)
0
5
10
15
20
NC1
NC2
BC1
BC2
D
E
F
UMI counts
0
10000
20000
30000
40000
50000
60000
70000
Genes per spot
0
1000
2000
3000
4000
5000
6000
7000
8000
Mitochondrial (%)
0.0
2.5
5.0
7.5
10.0
12.5
15.0
17.5
20.0
NC1
NC2
BC1
BC2

**Supplementary Figure 4 – Pre-filtering spatial transcriptomics (ST) UMI counts, genes per spot, and percent mitochondrial genes.**

**A-C** Whole tissue section spatial plots of pre-filtering unique molecular identifier (UMI) counts (**A**), genes per spot (**B**), and mitochondrial gene percentage (**C**). **D-F** Violin plots for pre-filtering UMI counts (**D**), genes per spot (**E**), and mitochondrial gene percentage (**F**). Violin plots show spot-level distributions; boxplots indicate median and interquartile range (IQR), with whiskers extending to 1.5 x IQR.

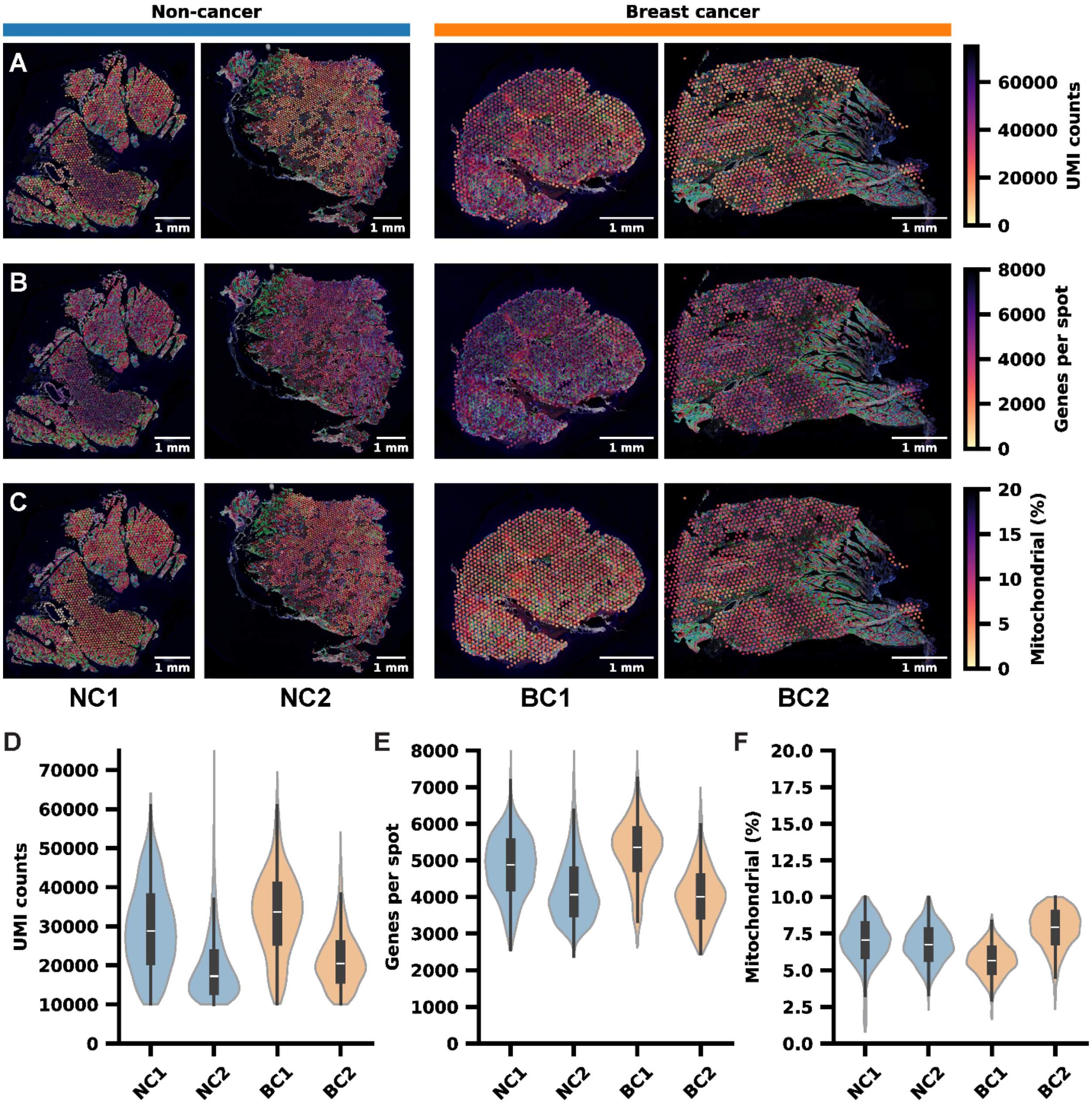

Non-cancer
Breast cancer
A
B
C
1 mm
UMI counts
60000
40000
20000
0
Genes per spot
8000
6000
4000
2000
0
Mitochondrial (%)
20
15
10
5
0
NC1
NC2
BC1
BC2
D
UMI counts
70000
60000
50000
40000
30000
20000
10000
0
E
Genes per spot
8000
7000
6000
5000
4000
3000
2000
1000
0
F
Mitochondrial (%)
20.0
17.5
15.0
12.5
10.0
7.5
5.0
2.5
0.0

**Supplementary Figure 5 – Post-filtering spatial transcriptomics (ST) UMI counts, genes per spot, and percent mitochondrial genes.**

**A-C** Whole tissue section spatial plots of post-filtering unique molecular identifier (UMI) counts (**A**), genes per spot (**B**), and mitochondrial gene percentage (**C**). **D-F** Violin plots for post-filtering UMI counts (**D**), genes per spot (**E**), and mitochondrial gene percentage (**F**). Violin plots show spot-level distributions; boxplots indicate median and interquartile range (IQR), with whiskers extending to 1.5 x IQR.

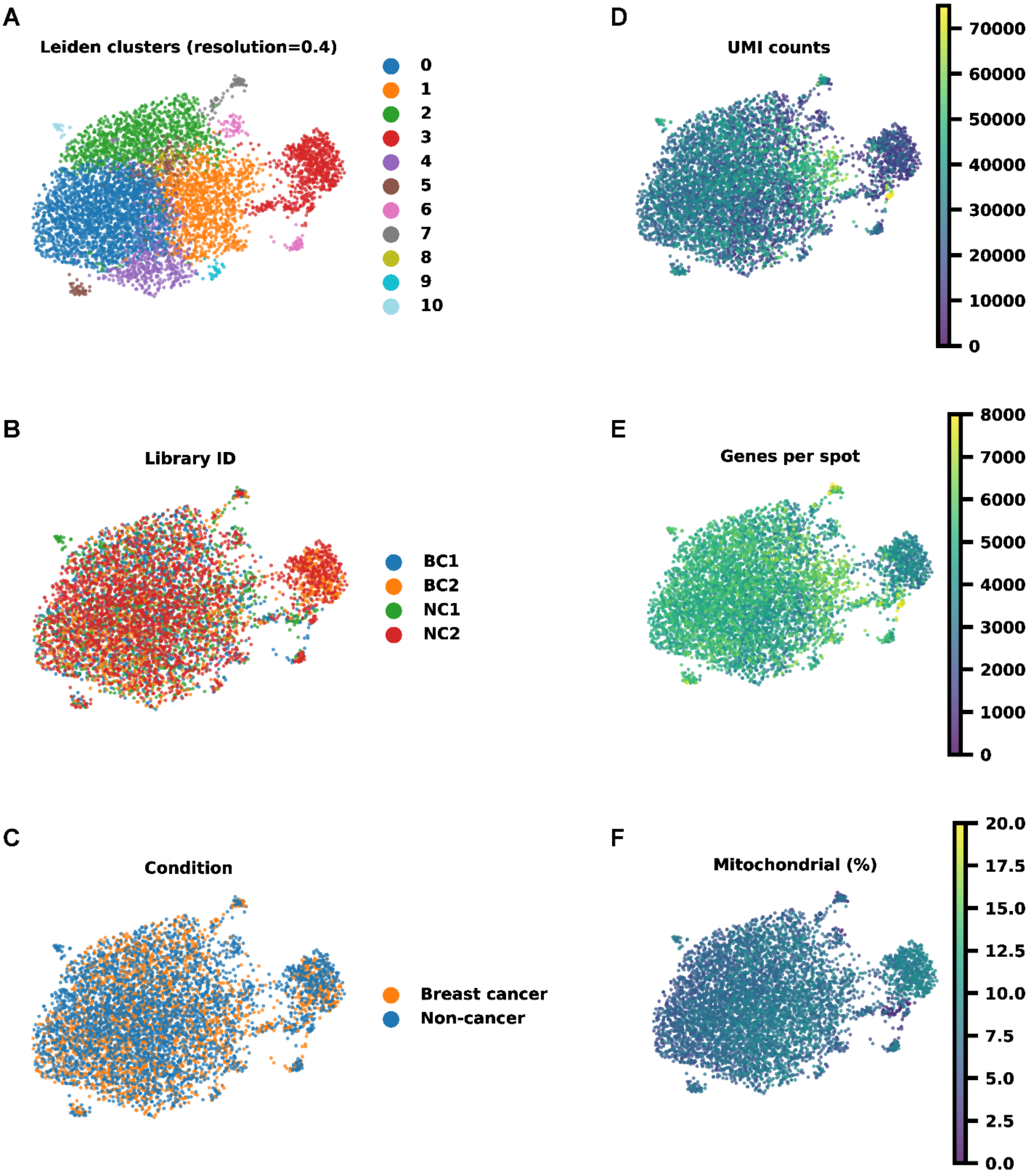
A
Leiden clusters (resolution=0.4)
0
1
2
3
4
5
6
7
8
9
10
D
UMI counts
70000
60000
50000
40000
30000
20000
10000
0
B
Library ID
BC1
BC2
NC1
NC2
E
Genes per spot
8000
7000
6000
5000
4000
3000
2000
1000
0
C
Condition
Breast cancer
Non-cancer
F
Mitochondrial (%)
20.0
17.5
15.0
12.5
10.0
7.5
5.0
2.5
0.0

**Supplementary Figure 6 – Batch correction and clustering quality control UMAPs.**

**A-F** Uniform manifold approximation and projection (UMAP) of spatial transcriptomic spots colored by Leiden cluster (resolution=0.4; **A**), library ID (**B**), study condition (non-cancer v. breast cancer; **C**), unique molecular identifier (UMI) counts (**D**), genes per spot (**E**), and mitochondrial gene percentage (**F**).
